# Time-dependent heterogeneity leads to transient suppression of the COVID-19 epidemic, not herd immunity

**DOI:** 10.1101/2020.07.26.20162420

**Authors:** Alexei V. Tkachenko, Sergei Maslov, Ahmed Elbanna, George N. Wong, Zachary J. Weiner, Nigel Goldenfeld

**Affiliations:** Center for Functional Nanomaterials, Brookhaven National Laboratory, Upton, NY 11973, USA; Department of Physics, University of Illinois at Urbana-Champaign, Urbana, IL 61801, USA; Department of Civil Engineering, University of Illinois at Urbana-Champaign, Urbana, IL 61801, USA; Department of Bioengineering, University of Illinois at Urbana-Champaign, Urbana, IL 61801, USA; Carl R. Woese Institute for Genomic Biology, University of Illinois at Urbana-Champaign, Urbana, IL 61801, USA

**Keywords:** COVID-19, Heterogeneity, Overdispersion, Epidemic theory

## Abstract

Epidemics generally spread through a succession of waves that reflect factors on multiple time-scales. On short time-scales, superspreading events lead to burstiness and overdispersion, while long-term persistent heterogeneity in susceptibility is expected to lead to a reduction in the infection peak and the herd immunity threshold (HIT). Here, we develop a general approach to encompass both time-scales, including time variations in individual social activity, and demonstrate how to incorporate them phenomenologically into a wide class of epidemiological models through parameterization. We derive a non-linear dependence of the effective reproduction number *R*_*e*_ on the susceptible population fraction *S*. We show that a state of transient collective immunity (TCI) emerges well below the HIT during early, high-paced stages of the epidemic. However, this is a fragile state that wanes over time due to changing levels of social activity, and so the infection peak is not an indication of herd immunity: subsequent waves can and will emerge due to behavioral changes in the population, driven (e.g.) by seasonal factors. Transient and long-term levels of heterogeneity are estimated by using empirical data from the COVID-19 epidemic as well as from real-life face-to-face contact networks. These results suggest that the hardest-hit areas, such as NYC, have achieved TCI following the first wave of the epidemic, but likely remain below the long-term HIT. Thus, in contrast to some previous claims, these reqions can still experience subsequent waves.

**Significance Statement**

Epidemics generally spread through a succession of waves that reflect factors on multiple time-scales. Here, we develop a general approach to encompass super-spreading and population heterogeneity, and demonstrate that a fragile state of transient collective immunity (TCI) emerges well below the HIT during early, high-paced stages of the epidemic. However, this is not an indication of herd immunity: subsequent waves can and will emerge due to behavioral changes in the population, driven (e.g.) by seasonal factors. Analysis of empirical data suggests that even in locations with strong first waves of COVID-19, subsequent waves will still emerge.

---

The COVID-19 pandemic is nearly unprecedented in the level of disruption it has caused globally, but also, potentially, in the degree to which it will change our understanding of epidemic dynamics and the efficacy of various mitigation strategies. Ever since the pioneering works of Kermack and McKendrick (1), epidemiological models have been widely and successfully used to quantify and predict the progression of infectious diseases (2–6). More recently, the important role in epidemic spreading played by population heterogeneity and the complex structure of social networks has been appreciated and highlighted in multiple studies (7–25). However, integration of this conceptual progress into reliable, predictive epidemiological models remains a formidable task. Among the key effects of heterogeneity and social network structure are (i) the role played by superspreaders and superspreading events during initial outbreaks (12, 13, 16, 26–28) and (ii) a substantial reduction of the final size of epidemic (FSE) as well as the herd immunity threshold (HIT) (9, 10, 19–21, 23, 29). The COVID-19 pandemic has re-ignited interest in the effects of heterogeneity of individual susceptibility to the disease, in particular to the possibility that it might lower both HIT and FSE (30–34). In studying epidemics in heterogeneous populations it is important to emphasize the qualitative nature of two important time-scales. First, overdispersion reflects short-term patterns of behavior and even accidental events, and not the degree of persistent population behavioral heterogeneity. Second, short-term overdispersion is generally supposed to have a limited impact on the long term epidemic dynamics, being important primarily in early outbreaks which are dominated by super-spreading events. In other words, epidemics are generally assumed to have distinct descriptions on widely-differing time-scales. In this paper, we attempt to provide a multi-scale theory for epidemic progression and show that both overdispersion and persistent heterogeneity affect the overall progression of the COVID-19 epidemic. The significance of this multi-scale perspective is that it provides a natural formalism to predict the occurrence and nature of successive epidemic waves, even when it might seem that a first wave has attained a state which could be mistaken for herd immunity.

There are several existing approaches to model the effects of heterogeneity on epidemic dynamics, each focusing on a different characteristic and parameterization. In the first approach, one stratifies the population into several demographic groups (e.g. by age), and accounts for variation in susceptibility of these groups and their mutual contact probabilities (2). While this approach represents many aspects of population dynamics beyond the homogeneous and well-mixed assumption, it clearly does not encompass the whole complexity of individual heterogeneity, interpersonal communications and spatial and social structures. These details can be addressed in a second approach, where one analyzes the epidemic dynamics on real-world or artificial social networks (8, 13, 23, 35, 36). Through elegant mathematics, it is possible to obtain detailed results in idealized cases, including the mapping onto well-understood models of statistical physics such as percolation (9). As demonstrated in Refs. (21) the FSE is a very robust property of the epidemic, insensitive to fine details of its dynamics but defined by (i) distribution of susceptibilities in the population (20, 29, 37); (ii) correlations between infectivity and susceptibility. Importantly, it does not depend on the part of heterogeneous infectivity that is not correlated with susceptibility. However, these approaches have so far been mostly limited to the analysis of the FSE and distribution of outbreak sizes on a static social network.

For practical purposes, it is desirable to predict the complete time-dependent dynamics of an epidemic, preferably by explicitly including heterogeneity into classical well-mixed mean-field compartmentalized models. This approach was developed some time ago in the context of epidemics on networks (10, 23) and the resulting mean-field theory effectively reproduces the structure of heterogeneous well-mixed models extensively studied in the applied mathematics literature (19–22, 24, 29). The overall impact of heterogeneity occurs because as the disease spreads, it preferentially “vaccinates” the more susceptible individuals, so the remaining population is less susceptible, and spread is inhibited. This effect is further enhanced by a positive correlation between infectivity and susceptibility. In the context of static network models this correlation is perfect since both factors are proportional to the degree of individual nodes. Ref. (24) studied a hybrid model in which social heterogeneity represented by network degree was combined with a biological one. These approaches have been recently applied in the context of COVID-19 (30, 31, 34, 38, 39). Here, the conclusion was that the herd immunity threshold may be well below that expected in classical homogeneous models.

These approaches to heterogeneity delineate end-members of a continuum of theories: overdispersion describing short-term, bursty dynamics (e.g. due to super-spreader accidents), as opposed to *persistent heterogeneity*, which is a long-term characteristic of an individual and reflects behavioral propensity to (e.g.) socialize in large gatherings without prudent social distancing. Overdispersion is usually modeled in terms of a negative binomial branching process (12, 13, 16, 26–28), and is expected to be a much stronger source of variation compared to the longer-term characteristics that reflect persistent heterogeneity. It is generally presumed that this short-term overdispersion has no effect on the epidemic dynamics outside the early outbreaks. Indeed, large variations in an individual’s infectivity would average out as long as they are not correlated with susceptibility. But since the initial exposure and secondary infections are separated by a single generation interval (typically about 5 days for COVID-19), the levels of individual social activity at those times are expected to be correlated, and (at least partially) reflect short-term overdispersion. How, then, can one understand the epidemic progression across various timescales, from early stages of a fast exponential growth to the final state of the herd immunity?

Below, we present a comprehensive yet simple theory that accounts for both social and biological aspects of heterogeneity, and predicts how together they modify early, and intermediate epidemic dynamics, as well as global characteristics of the epidemic such as its HIT. Importantly, early epidemic dynamics may be sensitive both to persistent heterogeneity and short-term overdispersion. As a result, the apparent early-stage heterogeneity is typically enhanced compared to its long-term persistent level. This may lead to a suppression of the first wave of the epidemic due to reaching a novel state that we call Transient Collective Immunity (TCI) determined by a combination of short-term and long-term heterogeneity, whose threshold is lower than the eventual HIT. The implication is that the first wave of an epidemic ends due to a combination of both persistent heterogeneity and whatever mitigation constraints are imposed on the population. As the latter are relaxed by authorities or through behavioral changes associated with seasonal factors, subsequent waves can still occur. Thus, TCI is dramatically different from the idea of herd immunity due to heterogeneity.

Our starting point is a generalized version of the heterogeneous well-mixed theory in the spirit of Ref. (10), but we use the age-of-infection approach (1) rather than compartmentalized SIR/SEIR models of epidemic dynamics (see, e.g. (2)). Similar to multiple previous studies, we first completely ignore any time dependence of individual susceptibilities and infectivities, focusing only on their long-term persistent components. This approach implicitly assumes that any short-term overdispersion (responsible, e.g. for the super-spreading phenomenon) is effectively averaged out. This is a valid assumption if the large short-term variations in individual infectivity are completely uncorrelated with an individual’s susceptibility. However, this approximation is hard to justify in the case of COVID-19. Indeed, if the two are correlated on the timescale of a single generation interval (5 days), this will strongly affect the overall epidemic dynamics. Therefore, our initial analysis is eventually expanded to a more general theory accounting for the non-negligible effects of short-term overdispersion. In the case of persistent heterogeneity we demonstrate how the model can be recast into an effective homogeneous theory that can readily encompass a wide class of epidemiological models, including various versions of the popular SIR/SEIR approaches. Specific innovations that emerge from our analysis are the non-linear dependence of the effective reproduction number *R*_*e*_ on the overall population fraction *S* of susceptible individuals, and another non-linear function *S*_*e*_ that gives an effective susceptible fraction, taking into account preferential removal of highly susceptible individuals.

A convenient and practically useful aspect of this approach is that it does not require extensive additional calibration in order to be applied to real data. In the effort to make quantitative predictions from epidemic models, accurate calibration is arguably the most difficult step, but is necessary due to the extreme instability of epidemic dynamics in both growth and decay phases (40, 41). We find that with our approach, the entire effect of heterogeneity is in many cases well-characterized by a single parameter which we call the *immunity factor λ*. It is related to the statistical properties of heterogeneous susceptibility across the population and to its correlation with individual infectivity. The immunity factor determines the rate at which *R*_*e*_ drops during the early stages of the epidemic as the pool of susceptibles is being depleted: *R*_*e*_ ≈*R*_0_(1− *λ*(1 − *S*)). Beyond this early linear regime, for an important case of gamma-distributed individual susceptibilities, we show that the classical proportionality, *R*_*e*_ = *R*_0_*S*, transforms into a power-law scaling relationship *R*_*e*_ = *R*_0_*S*^*λ*^. This leads to a modified version of the result for the herd immunity threshold, 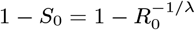.

Heterogeneity in the susceptibility of individual members of the population has several different contributions: (i) biological, which takes into account differences in factors such as strength of immune response, genetics, age, and comorbidities; and (ii) social, reflecting differences in the number and frequency of close contacts of different people. The immunity factor *λ* in our model combines these sources of heterogeneous susceptibility as well as its correlation with individual infectivity. As we demonstrate, under certain assumptions the immunity factor is simply a product of social and biological contributions: *λ* = *λ*_*s*_*λ*_*b*_. In our study, we leverage existing studies of real-life face-to-face contact networks (13, 19, 36, 42– 45) to estimate the social contribution to heterogeneous susceptibility, and the corresponding immunity factor *λ*_*s*_. The biological contribution, *λ*_*b*_, is expected to depend on specific details of each infection. For the case of COVID-19, there is little indication that biological variations in susceptibility, unrelated to one’s social activity, play a significant role in the epidemic dynamics.

To test this theory, we use empirical data for the COVID-19 epidemic to independently estimate the immunity factor *λ*. In particular, we apply our previously-described epidemic model that features multi-channel Bayesian calibration (40) to describe epidemic dynamics in New York City and Chicago. This model uses high quality data on hospitalizations, Intensive Care Unit (ICU) occupancy and daily deaths to extract the underlying *R*_*e*_(*S*) dependence in each of two cities. In addition, we perform a similar analysis of data on individual states in the USA, using data generated by the model in Ref. (46). Using both approaches, we find that the locations that were severely impacted by the COVID-19 epidemic show a more pronounced reduction of the effective reproduction number. This effect is much stronger than predicted by classical homogeneous models, suggesting a significant role of heterogeneity. The estimated immunity factor ranges between 4 and 5. Importantly, this represents a transient value of the parameter *λ* observed on intermediate timescales and dependent on both persistent and short-term heterogeneity. Our estimates of the long-term, value of the immunity factor, defined by persistent heterogeneity only, is considerably lower: *λ*_∞_≃2. This difference explain why achieving the state of transient collective immunity after the first wave of the epidemic does not imply long-term herd immunity.

Finally, we integrate the persistent heterogeneity theory into our time-of-infection epidemiological model (40), and project possible outcomes of the second wave of the COVID-19 epidemic in during the summer months in NYC and Chicago, using data up to the end of May 2020. By considering the worst-case scenario of a full relaxation of any currently imposed mitigation, we find that the results of the heterogeneity-modified model significantly modify the results from the homogeneous mode. In particular, based on our estimate of the immunity factor, we expect virtually no second wave in NYC in the immediate future, indicating that the TCI has likely been achieved there. Chicago, on the other hand, has not passed the TCI threshold that we infer, but the effects of heterogeneity would still result in a substantial reduction of the magnitude of the second wave there, even under the worst-case scenario.

This, in turn, suggests that the second wave can be completely eliminated in such medium-hit locations, if appropriate and economically mild mitigation measures are adopted, including e.g. mask wearing, contact tracing, and targeted limitation of potential super-spreading events, through limitations on indoor bars, dining and other venues. We further investigate the issue of fragility of collective immunity in heterogeneous populations. By allowing rewiring of the social network with time, we demonstrate that the TCI may wane, much like an individual’s acquired immunity may wane due to biological factors. This phenomenon would result in the emergence of secondary epidemic waves after a substantial period of low infection counts.

## Theory of epidemics in populations with persistent heterogeneity

Following in the footsteps of Refs.(10, 19, 22–24, 29, 31), we consider the spread of an epidemic in a population of individuals who exhibit significant persistent heterogeneity in their susceptibilities to infection *α*. This heterogeneity may be biological or social in origin, and we assume these factors are independent: *α* = *α*_*b*_ *α*_*s*_. Effects of possible correlations between *α*_*b*_ and *α*_*s*_ have been discussed in Ref. (24). The biologically-driven heterogeneous susceptibility *α*_*b*_ is shaped by variations of several intrinsic factors such as the strength of individuals’ immune responses, age, or genetics. In contrast, the socially-driven heterogeneous susceptibility *α*_*s*_ is shaped by extrinsic factors, such as differences in individuals’ social interaction patterns (their degree in the network of social interactions). Furthermore, individuals’ different risk perceptions and attitudes towards social distancing may further amplify variations in socially-driven susceptibility heterogeneity. We only focus on susceptibility that is a persistent property of an individual. For example, people who have elevated occupational hazards, such as healthcare workers, typically have higher, steady values of *α*_*s*_. Similarly, people with low immune response, highly social individuals (hubs in social networks), or scofflaws would all be characterized by above-average overall susceptibility *α*.

In this work, we group individuals into sub-populations with similar values of *α* and describe the heterogeneity of the overall population by the probability density function (pdf) of this parameter, *f* (*α*). Since *α* is a relative measure of individual susceptibilities, without loss of generality we Set 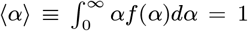. Each person is also assigned an individual reproduction number *R*_*i*_, which is an expected number of people that this person would infect in a fully susceptible population with ⟨*α*⟩ = 1. Accordingly, from each sub-population with susceptibility *α* there is a respective mean infectivity or reproductive number *R*_*α*_. Any correlations between individual susceptibility and infectivity will significantly impact the epidemic dynamics. Such correlations are an integral part of most network-based epidemiological models due to the assumed reciprocity in underlying social interactions, which leads to *R*_*α*_∼ *α* (9, 10, 23). In reality, not all transmissions involve face-to-face contacts, and biological susceptibility need not be strongly correlated with infectivity. Therefore, it is reasonable to expect only a partial correlation between *α* and *R*_*α*_.

Let *S*_*α*_(*t*) be the fraction of susceptible individuals in the subpopulation with susceptibility *α*, and let 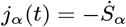 be the corresponding daily incidence rate, i.e., the fraction of newly infected individuals per day in that sub-population. At the start of the epidemic, we assume everyone is susceptible to infection: *S*_*α*_(0) = 1. The course of the epidemic is described by the following age-of-infection model:

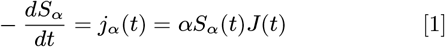

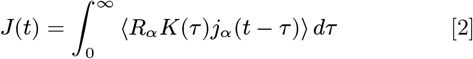

Here *t* is the physical time and *τ* is the time since infection for an individual.⟨…⟩ represents averaging over *α* with pdf *f* (*α*). *J* (*t*) represents the mean daily attack rate, i.e. hypothetical incidence rate in a fully susceptible homogeneous population with *α* = 1. *K*(*τ*) is the distribution of the generation interval, which we assume to be independent of *α* for the sake of simplicity.

According to Eq. (1), the susceptible subpopulation for any *α* can be expressed as

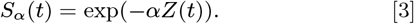

Here 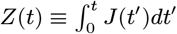. The total susceptible fraction of the population is related to the moment generating function *M* _*α*_ of the distribution *f* (*α*) (i.e., the Laplace transform of *f* (*α*)) according to:

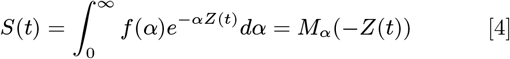

Similarly, the effective reproductive number *R*_*e*_ can be expressed in terms of the parameter *Z*:

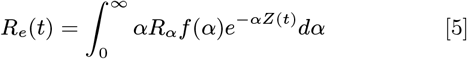

Note that for *Z* = 0, this expression gives the basic reproduction number *R*_0_ = ⟨ *αR*_*α*_⟩. Since both *S* and *R*_*e*_ depend on time only through *Z*(*t*), Eqs. (4)–(5) establish a parametric relationship between these two important quantities during the time course of an epidemic. In contrast to the classical case when these two quantities are simply proportional to each other, i.e. *R*_*e*_ = *SR*_0_, the relationship in the present theory is non-linear due to heterogeneity. Now one can re-write the renewal equation for the force of infection in the same form as if this were a homogeneous problem:

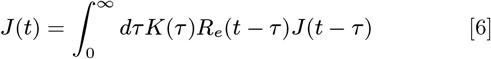

Furthermore, by averaging Eq. (1) over all values of *α* one arrives at the following heterogeneity-induced modification to the relationship between the force of infection and incidence rate:

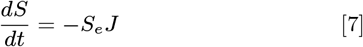

Here

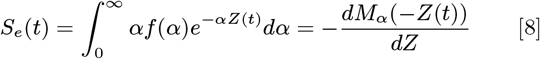

is the effective susceptible fraction of the population, which is less than *S* due to the disproportionate removal of highly susceptible individuals. Just as with *R*_*e*_, it is a non-linear function of *S*, defined parametrically by Eqs. (4,8). Further generalization of this theory for the time-modulated age-of-infection model is presented in the *Supporting Information Appendix (SI Appendix)*. There, we also discuss the adaptation of this approach for the important special case of a compartmentalized SIR/SEIR model. Such non-linear modifications to homogeneous epidemiological models have been proposed in the past, both as plausible descriptions of heterogeneous populations and in other contexts (17–19). However, those empirical models exhibited a limited range of applicability (19) and have not had a solid mechanistic foundation, with a noticeable exception of a special case of SIR model without correlation between susceptibility and infectivity studied in Ref. (29). Our approach is more general: it provides the exact mapping of a wide class of heterogeneous well-mixed models onto homogeneous ones, and provides a specific relationship between the underlying statistics of *α* and *R*_*α*_ and the non-linear functions *R*_*e*_(*S*) and *S*_*e*_(*S*).

One of the striking consequences of the non-linearity of *R*_*e*_(*S*) is that the effective reproduction number could be decreasing at the early stages of an epidemic significantly faster than predicted by homogeneous models. Specifically, for 1 − *S*(*t*) ≃ *Z*(*t*) <<1 one can linearize the effective reproduction number as

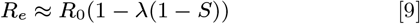

We named the coefficient *λ* the *immunity factor* because it quantifies the effect that a gradual build-up of population immunity has on the spread of an epidemic. The classical value of *λ* is 1, but it may be significantly larger in a heterogeneous case. By linearizing Eq. (5) in terms of 1 − *S* ≃ *Z* ≪ 1 and dividing the result by *R*_0_ = ⟨*αR*_*α*_⟩ one gets:

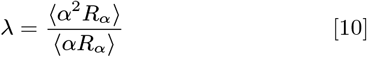

As one can see, the value of the immunity factor, thus depends both on the statistics of susceptibility *α*, and on its correlation with infectivity *R*_*α*_.

We previously defined the overall susceptibility as a combination of biological and social factors: *α* = *α*_*s*_*α*_*b*_ Here *α*_*s*_ is a measure of the overall social connectivity or activity of an individual, such as the cumulative time of close contact with other individuals averaged over a sufficiently long time interval (known as node strength in network science). Since the contribution of interpersonal contacts to an epidemic spread is mostly reciprocal, we assume *R*_*α*_ ∼*α*_*s*_. On the other hand, in our analysis we neglect a correlation between biological susceptibility and infectivity, as well as between *α*_*b*_ and *α*_*s*_. Under these approximations, the immunity factor itself is a product of biological and social contributions, *λ* = *λ*_*b*_*λ*_*s*_. Each of them can be expressed in terms of leading moments of *α*_*b*_ and *α*_*s*_, respectively:

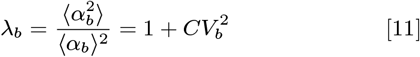

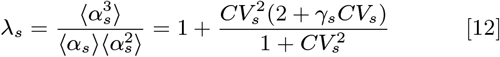

These equations follow from Eq. 10 in the limit *R*_*α*_ = const and *R*_*α*_ ∼*α* respectively. Although these equations resemble classical results for *R*_0_ in heterogeneous networks (8–10, 23), here they describe a completely different effect of suppression of *R*_*e*_ in response to depletion of susceptible population *S*. That is why *λ*_*s*_ in Eq. 12 is proportional to the third moment of *α*_*s*_ instead of the second moment in the case of 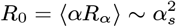. Note that the biological contribution to the immunity factor depends only on the coefficient of variation *CV*_*b*_ of *α*_*b*_. On the other hand, the social factor *λ*_*s*_ depends both on the coefficient of variation *CV*_*s*_ and the skewness *γ*_*s*_ of the distribution of *α*_*s*_. Due to our normalization, ⟨*α*_*s*_⟩ ⟨*α*_*b*_⟩ ≈ ⟨ *α*_*s*_*α*_*b*_⟩ = ⟨ *α* ⟩= 1.

The relative importance of biological and social contributions to the overall heterogeneity of *α* may be characterized by a single parameter *χ*. For a log-normal distribution of *α*_*b*_, *χ* appears as a scaling exponent between infectivity and susceptibility: *R*_*α*_∼ *α*^*χ*^ (see *SI Appendix* for details). The corresponding expression for the overall immunity factor is *λ* = ⟨*α*^2+*χ*^⟩ */ ⟨α*^1+*χ*^⟩. The limit *χ* = 0 corresponds to a predominantly biological source of heterogeneity, i.e., 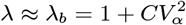, where *CV*_*α*_ is the coefficient of variation for the overall susceptibility. In the opposite limit *χ* = 1, population heterogeneity is primarily of social origin, hence *λ ≈λ*_*s*_ is affected by both *CV*_*α*_ and the skewness *γ*_*α*_ of the pdf *f* (*α*). The biological contribution *λ*_*b*_ depends on specific biological details of the disease and thus is unlikely to be as universal and robust as the social one. For the COVID-19 epidemic, there is no strong evidence of a wide variation in attack rates unrelated to social activity, geographic location, or socioeconomic status. For instance, there is very little age variability in COVID-19 prevalence as reported by the NYC Department of Health (47) based on the serological survey that followed the first wave of the epidemic. Therefore, below we will largely ignore possible biological heterogeneity, and focus on social heterogeneity.

So far, our discussion has focused on the early stages of epidemics, when the *R*_*e*_(*S*) dependence is given by a linearized expression Eq. (9). To describe the non-linear regime, we consider a gamma-distributed susceptibility: *f*(*α*) ∼ *α*^1/*η*−1^exp(-*α*/*η*),where 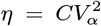. In this case, according to Eqs. (4) and (5), *R*_*e*_, *S*_*e*_ and *S* are related by scaling relationships (see *SI Appendix*):

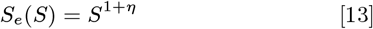

and

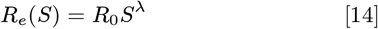

The exponent 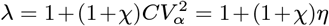 coincides with the early-epidemics immunity factor defined in Eqs. (9)–(10) for a general case. Note that without correlation (*χ* = 0), both scaling exponents would be the same; this result has been previously obtained for the SIR model in Ref. (29) and more recently reproduced in Ref. (38) in the context of COVID-19. The scaling behavior *R*_*e*_(*S*) is shown in Fig. 1 for *λ* = 3 ± 1. This function is dramatically different from the classical linear dependence *R*_*e*_ = *SR*_0_. To emphasize the importance of this difference, we indicate the estimated fractions of the population in New York City and Chicago susceptible to COVID-19, as of the end of May 2020.

**Fig. 1.**
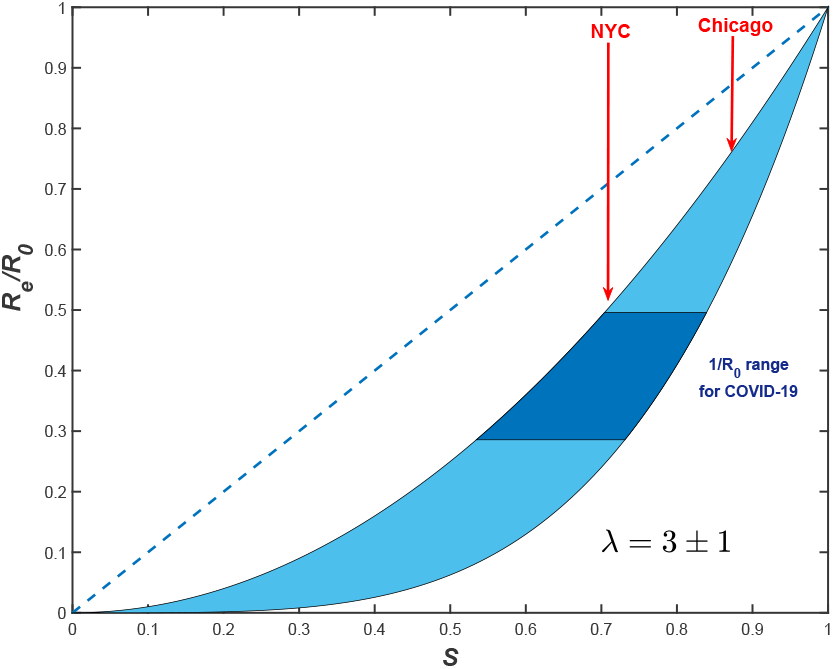
*R*_*e*_*/R* vs *S* dependence for gamma-distributed susceptibility with *λ* = 3*±*1 (blue area). The dashed line shows the classical homogeneous result, *R*_*e*_ = *R*_0_ *S*. Note a substantial reduction of *R*_*e*_ for COVID-19 epidemic expected in both NYC and Chicago, compared to that value. Approximate fractions of susceptible populations, *S*,for both cities are estimated as of the end of May 2020, by using the model described in Ref. (40).

Eq. (14) immediately leads to a major revision of the classical result for the herd immunity threshold 1−*S*_0_ = 1 − 1*/R*_0_. *S*_0_ is the fraction of susceptible population at which the exponential growth stops, while 1 − *S*_0_ is the relative size of the epidemic at that time. By setting *R*_*e*_ = 1 in Eq. (14), we obtain:

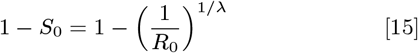

As we were finalizing this paper for public release, a preprint by Aguas et al. appeared online (48) that independently obtained our Eqs. (14, 15) for gamma-distributed susceptibilities. The same result has also been recently obtained in Ref. (39). Note however that the full quasi-homogeneous description of the epidemic dynamics requires both *R*_*e*_(*S*) and *S*_*e*_(*S*), that in the general case is characterized by different scaling exponents.

Our focus on the gamma distribution is well justified by the observation that the social strength *α*_*s*_ is approximately exponentially distributed, i.e., it is a specific case of the gamma distribution with 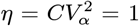 (see more discussion of this in the next section). A moderate biological heterogeneity would lead to an increase of the overall *CV*_*α*_, but the pdf *f* (*α*) will still be close to the gamma distribution family. From the conceptual point of view, it is nevertheless important to understand how the function *R*_*e*_(*S*) would change if *f* (*α*) had a different functional form. In *SI Appendix*, we present analytic and numerical calculations for two other families of distributions: (i) an exponentially bounded power law *f* (*α*) ∼*e*^−*α/α*+^ */α*^*q*^ (*q*≥ 1, with an additional cut-off at lower values of *α*) and (ii) the log-normal distribution. In addition, we give an approximate analytic result that generalizes Eq. (14) for an arbitrary skewness of *f* (*α*). This generalization works remarkably well for all three families of distributions analyzed in this work. As suggested by Eqs. (11-12), as the distribution becomes more skewed, the range between the *χ* = 0 and *χ* = 1 curves broadens. For instance, for distributions dominated by a power law, *f* (*α*) ∼1*/α*^*q*^, with 3 *< q <* 4 and *χ* = 1, *λ* diverges even though *CV*_*α*_ remains finite. This represents a crossover to the regime of so-called scale-free networks (2 ≤ *q* ≤3), which are characterized by zero epidemic threshold yet strongly self-limited dynamics: the epidemic effectively kills itself by immunizing the hubs on the network (10, 23, 49).

## Role of short-term variations in social activity

Short-term overdispersion in transmission is commonly presumed to have no effect on the overall epidemic dynamics, aside from the early outbreak often dominated by superspreaders. This would indeed be the case if overdispersed transmission were completely uncorrelated with individual susceptibility. But since the timescale for an individual’s infectivity (about 2 days) is comparable to a single generation interval (about 5 days) for the COVID-19 epidemics, ignoring such correlations appears unreasonable. We therefore developed a generalization of the theory presented in the previous section, that takes into account a time dependence of individual susceptibilities and infectivities, as well as temporal correlations between them. The theory is presented using a path-integral formulation in *SI Appendix*. Here we present several important results directly related to the transient suppression of an epidemic and differentiate these effects from herd immunity.

Since fast variations are primarily caused by bursty dynamics of social interactions (50–53), and since heterogeneous biological susceptibility appears subdominant in the context of COVID-19, we set *α*_*b*_ = 1 for all individuals. So *α* has purely social origin. Let *a*_*i*_(*t*) = *α*_*i*_ + *δa*_*i*_(*t*) be the time-dependent susceptibility of a person, which we associate with variable level of social activity. Hence, the same function determines also individual infectivity *a*(*t*)*R* around time *t*. Interestingly,even the classical result for basic reproduction number in a heterogeneous system, = *R*_0_ *R*⟨*α*^2^⟩, needs to be modified due to correlated short-term variations in social activity:

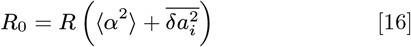

In the time-dependent generalization of our theory *Re* and *S* no longer have a fixed functional relationship between them. Instead, this relationship becomes non-local in time. For instance, our result for the suppression of *Re* at the early stages of the epidemic is still formally valid, but the effective value of immunity factor *λ* becomes time dependent, and Eqs.(9)-(10) become

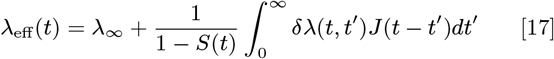

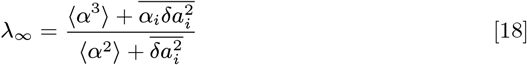

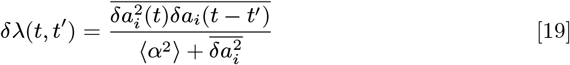

Constant *λ*_*∞*_ reflects suppression of *R*_*e*_ due to the build-up of the long-term collective immunity. On the other hand, the time-dependent term *δλ*(*t*) leads to an additional suppression of *R*_*e*_ over intermediate timescales. This term has likely played a significant role in shaping the transient self-limiting dynamics during the first wave of COVID-19 epidemic in some hard-hit locations.

Note that according to Eq. (17), *δλ*(*t‘*) is being averaged with the weight proportional to the force of infection *J* (*t* − *t*^*′*^) since 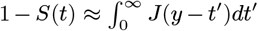 Since *δλ*(*t, t*^*′*^) decreases with time difference *t*^*′*^, its effect on *λ*_eff_ should be the strongest during the initial period of fast exponential growth. The initial suppression of the epidemic is caused by the combined effect of mitigation measures and both terms in *λ*_eff_. Since *λ*_eff_ *> λ*_*∞*_, the population may reach the state of Transient Collective Immunity (TCI) earlier than the actual long term Herd Immunity determined by persistent heterogeneity. However, this state is fragile and may wane with time. Specifically, as *J* (*t*) drops after the first wave, the second term in Eq. (17) gradually decays, bring *λ*_eff_ (*t*) closer to *λ*_*∞*_. According to Eq.(19), it is the correlation time of bursty social activity *δa*(*t*) that sets the timescale over which this TCI state deteriorates, and the new epidemic wave may get ignited. The relationship between this relaxation time and the duration of a single epidemic wave also determines the typical value of *λ*_eff_ during that wave.

Despite a large number of empirical studies of contact networks (50–53), information about the temporal correlations in *α*(*t*) or its proxies remains limited. On the other hand, much more is known about parameters of persistent heterogeneity. Recently, real-world networks of face-to-face communications have been studied using a variety of tools, including RFID devices (42), Bluetooth and Wi-Fi wearable tags, smartphone apps (43, 44), as well as census data and personal surveys (13, 36, 45). Despite coming from a wide variety of contexts, the major features of contact networks are remarkably robust. In particular, pdfs of both the degree (the number of contacts per person), and the node strength plotted in log-log coordinates appear nearly constant followed by a sharp fall after a certain upper cut-off. This behavior is generally consistent with an exponential distribution in *f*_*s*_(*α*_*s*_) (19, 43, 45), *f* (*α*) ∼ *e*^−*α/*⟨*α*⟩^.That sets the value of 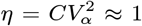. If not for short-term overdispersion, that would yield 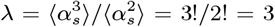 according to Eq. (12). However, with temporal effects taken into account, the build-up of long-term collective immunity is determined by *λ*_eff_ (*∞*) = *λ*_*∞*_. In order to estimate it, we make a simple model assumption that the short-term overdispersion for a particular individual is proportional to the persistent value of that person’s social activity: 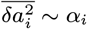. This leads to

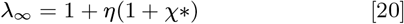

Here 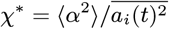 is a parameter that measures the relative strength of persistent heterogeneity and the overdispersion on the timescale of a single generation interval. Note that formally we recover our original result for *λ* in the purely persistent case, with *χ*^*∗*^ replacing the parameter *χ* that originally quantified the correlation between infectivity and susceptibility. By assuming the limit of strong short-term overdispersion (*χ*^*∗*^*<<* 1), we obtain *λ*_*∞*_ ≈ 2. As shown in *SI Appendix*, the very same value of *λ*_*∞*_ should be used as a scaling exponent for long-term behavior of *R*_*e*_(*S*). Therefore, HIT is set by Eq.(15) with *λ* = *λ*_∞_ ≈ 2. Its value is plotted vs. *R*_0_ in Fig. 2, along with the homogeneous result, and the estimated threshold of *TCI*. To estimate the corresponding transient immunity factor *λ*_eff_, we analyze the empirical data for the first wave of the COVID-19 epidemic below.

**Fig. 2.**
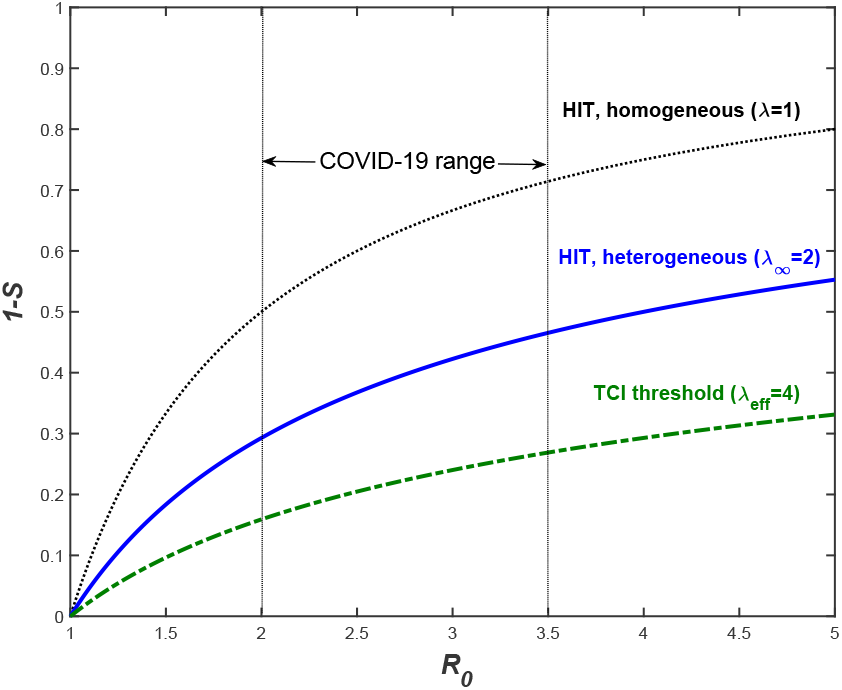
TCI threshold (dot-dashed), long-term heterogeneous HIT (solid), and homogeneous HIT (dotted) for various values of *R*_0_ (x-axis). HIT (solid line) is determined by persistent heterogeneity. The corresponding immunity factor *λ*_*∞*_ ≈ 2 is estimated from empirical face-to-face contact network. For transient behavior *λ*_eff_ ≈ 4 is assumed based on analysis of empirical data for COVID-19 epidemic in select locations.

## Application to the COVID-19 epidemic

The COVID-19 epidemic reached the US in early 2020, and by March it was rapidly spreading across multiple states. The early dynamics was characterized by a rapid rise in the number of cases with doubling times as low as 2 days. In response to this, the majority of states imposed a broad range of mitigation measures including school closures, limits on public gatherings, and Stay-at-Home orders. In many regions, especially the hardest-hit ones like New York City, people started to practice some degree of social distancing even before government-mandated mitigation. In order to quantify the effects of heterogeneity on the spread of the COVID-19 epidemic, we apply the Bayesian age-of-infection model described in Ref. (40) to New York City and Chicago. For both cities, we have access to reliable time series data on hospitalization, ICU room occupancy, and daily deaths due to COVID-19 (47, 54–56). We used these data to perform multi-channel calibration of our model (40), which allows us to infer the underlying time progression of both *S*(*t*) and *R*_*e*_(*t*). The fits for *R*_*e*_(*S*) for both cities are shown in Fig. 3A. In both cases, a sharp drop of *R*_*e*_ that occurred during the early stage of the epidemic is followed by a more gradual decline. For NYC, there is an extended range over which *R*_*e*_(*S*) has a constant slope in logarithmic coordinate. This is consistent with the power law behavior predicted by Eq. 14 with the slope corresponding to transient immunity factor *λ*_eff_ = 4.5 ± 0.05. Chicago exhibits a similar behavior but over a substantially narrower range of *S*. This reflects the fact that NYC was much harder hit by the COVID-19 epidemic. Importantly, the range of dates we used to estimate the immunity factor corresponds to the time interval after state-mandated Stay-At-Home orders were imposed, and before the mitigation measures began to be gradually relaxed. The signatures of the onset of the mitigation and of its partial relaxation are clearly visible on both ends of the constant-slope regime. To examine the possible effects of variable levels of mitigation on our estimates of *λ*_eff_ in Fig. S2 we repeated our analysis in which *R*_*e*_(*t*) was corrected by Google’s community mobility report in these two cities (57) (see *SI Appendix*). Although the range of data consistent with the constant slope shrank somewhat, our main conclusion remains unchanged. This provided us with a lower bound estimate for the transient immunity factor: *λ*_eff_ = 4.1±0.1.

**Fig. 3.**
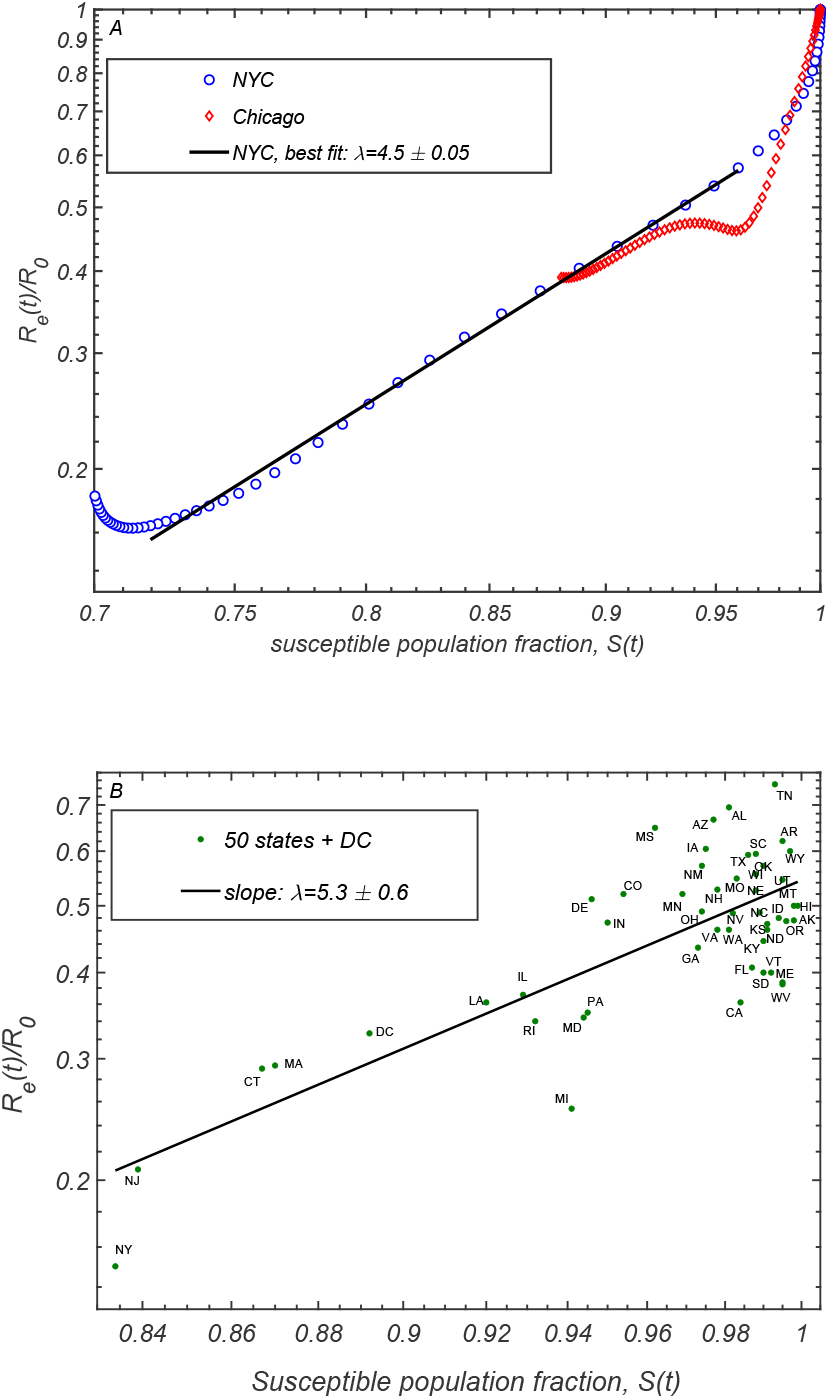
Correlation between the relative reduction in the effective reproduction number *R*_*e*_(*t*)*/R*_0_ (y-axis) with the susceptible population *S*(*t*). In Panel A, we present the progression of these two quantities for New York City and Chicago, as given by the epidemiological model described in Ref. (40). Panel B shows the scatter plot of *R*_*e*_(*t*_0_)*/R*_0_ and *S*(*t*_0_) in individual states of the US, evaluated in Ref. (46) (*t*_0_ is the latest date covered in that study).

To test the sensitivity of our results to details of the epidemiological model and choice of the region we performed an alternative analysis based on the data reported in (46). In that study, the COVID-19 epidemic was modelled in each of the 50 US states and the District of Columbia. Because of the differences in population density, level of urbanization, use of public transport, etc., different states were characterized by substantially different initial growth rates of the epidemic, as quantified by the basic reproduction number *R*_0_. Furthermore, the time of arrival of the epidemic also varied a great deal between individual states, with states hosting major airline transportation hubs being among the earliest ones hit by the virus. As a result of these differences, at any given time the infected fraction of the population differed significantly across the US (46). We use state level estimates of *R*_*e*_(*t*), *R*_0_ and *S*(*t*) as reported in Ref. (46) to construct the scatter plot *R*_*e*_(*t*_0_)*/R*_0_ vs *S*(*t*_0_) shown in 3, with *t*_0_ chosen to be the last reported date in that study, May 17, 2020. By performing the linear regression on these data in logarithmic coordinates, we obtain the fit for the slope *λ*_eff_ = 5.3 ± 0.6 and for *S* = 1 intercept around 0.54. In Fig. S3 (see *SI Appendix*), we present an extended version of this analysis for the 10 hardest-hit states and the District of Columbia, which takes into account the overall time progression of *R*_*e*_(*t*) and *S*(*t*), and gives similar estimate *λ*_eff_ = 4.7 *±* 1.5. Both estimates of the immunity factor based on the state data are consistent with our earlier analysis of NYC and Chicago. In light of our theoretical picture, this value of this transient immunity factor, *λ*_eff_ ≃ 4, is set by the the pace of the first epidemic wave in the US. As expected, it exceeds our estimate of *λ*∞ ≈2 associated with persistent heterogeneity and responsible for the long-term herd immunity.

We can now incorporate this transient level of heterogeneity into our epidemiological model, and examine how future projections change as a result of this modification. This is done by plugging scaling relationships given by Eqs. (13)-(14) into the force of infection and incidence rate equations of the original model. These equations are similar to Eqs. (6)-(7), but also include time modulation due to the mitigation and a possible seasonal forcing (see *SI Appendix* for more details). After calibrating the model by using the data streams on ICU occupancy, hospitalization and daily deaths up to the end of May, we explore a hypothetical worst-case scenario in which any mitigation is completely relaxed as of June 1, in both Chicago and NYC. In other words, the basic reproduction number *R*_0_ is set back to its value at the initial stage of the epidemic, and the only factor limiting the second wave is the partial or full TCI, *R*_*e*_ = *R*_0_*S*^*λ*^. The projected daily deaths for each of the two cities under this (unrealistically harsh) scenario are presented in Fig. 4 for various values of *λ*. For both cities, the homogeneous model (*λ* = 1, blue lines) predicts a second wave which is larger than the first one with an additional death toll of around 35, 000 in NYC and 12, 800 in Chicago. The magnitude of the second wave is greatly reduced by heterogeneity, resulting in no second wave in either of the two cities for *λ* = 5 (black lines). Even for a modest value *λ* = 3 (red lines), which is less than our estimate, the second wave is dramatically reduced in both NYC and Chicago (by about 90% and 70%, respectively).

**Fig. 4.**
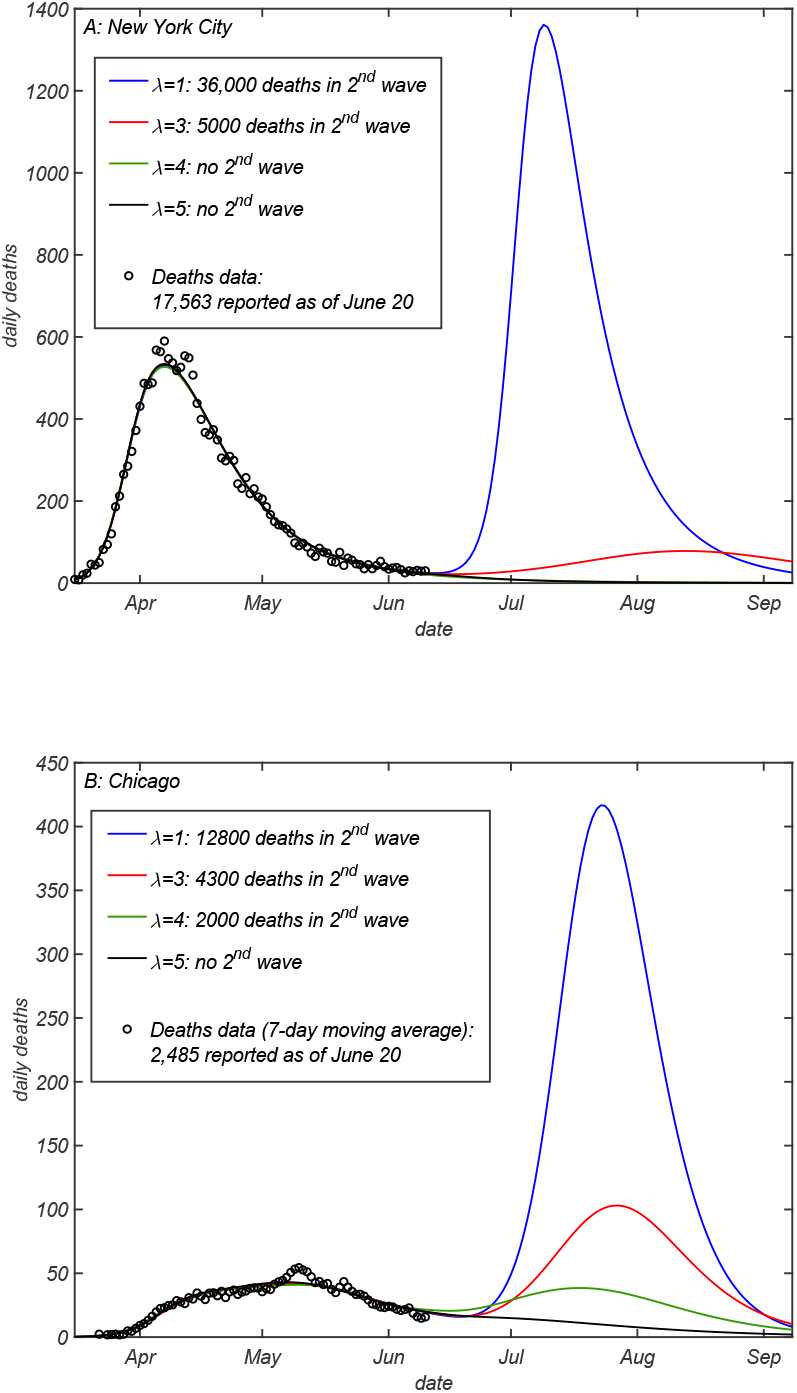
Projections of daily deaths under the hypothetical scenario in which any mitigation is completely eliminated as of June 15 2020, for (A) NYC and (B) Chicago. Different curves correspond to different values of the transient immunity factor *λ*_eff_ =1 (blue), 3 (red), 4 (green) and 5 (black lines). The model described in Ref. (40) was fully calibrated on daily deaths (circles), ICU occupancy and hospitalization data up to the end of May. See *SI Appendix*, Figs. S4-S5 in for additional details, including confidence intervals.

## Fragility of Transient Collective Immunity

One of the consequences of the bursty nature of social interactions is that the state of TCI gradually wanes due to changes of individual social interaction patterns on timescales longer than single generation interval. This may be viewed as a slow rewiring of social networks. In the context of the COVID-19 epidemic, individual responses to mitigation factors such as Stay-at-Home orders may differ across the population. When mitigation measures are relaxed, individual social susceptibility *α*_*s*_ inevitably changes. The impact of these changes on collective immunity depends on whether each person’s *α*_*s*_ during and after the mitigation are sufficiently correlated. For example, the TCI state would be compromised if people who practiced strict self-isolation compensated for it by an above-average social activity after the first wave of the epidemic has passed.

To illustrate the effects of post-mitigation rewiring of social networks we consider a simple modification of the heterogeneous model with no persistent heterogeneity (*α* = 1 for everyone) and exponentially distributed instantaneous levels of social activity *a*_*i*_(*t*). This corresponds to *λ*_eff_ (0) = 3 and *λ*_eff_(∞) = *λ*_∞_ = 1. In this model each individual completely changes the set of his/her social connections at some time scale *τ*_*s*_. These changes *destroy heterogeneity* giving rise to gradual relaxation of susceptible fraction *S*_*a*_ towards its overall mean value *S*. To model this, we modify Eq. 1 to include a simple relaxation term:

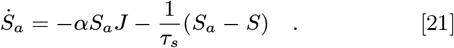

Epidemiological models with rewiring of underlying social networks have been studied before (58) (see (59) for a review), but under a constraint that the individual level of social activity quantified by network degree is preserved. In contrast, the dynamics described above stems from the individual level of social activity *α*_*s*_ changing in time.

Fig. 5 shows the simulation of SIR model, where the first wave of the epidemic is mitigated, thereby reducing the effective reproduction number *R*_0_ = 2.5. During the course of the mitigation *R*_0_ is multiplied by *µ* = 0.7. After the mitigation measures have been lifted at the end of the first wave, the population is positioned slightly below the TCI threshold preventing the immediate start of the second wave. However, gradual rewiring of the social network with time constant *τ*_*s*_ = 150 days ultimately results in the second and even the third wave of the epidemic (see Fig. 5). The inset of this figure shows *R*_*e*_(*t*) plotted as a function of *S*(*t*) in this epidemic.

**Fig. 5.**
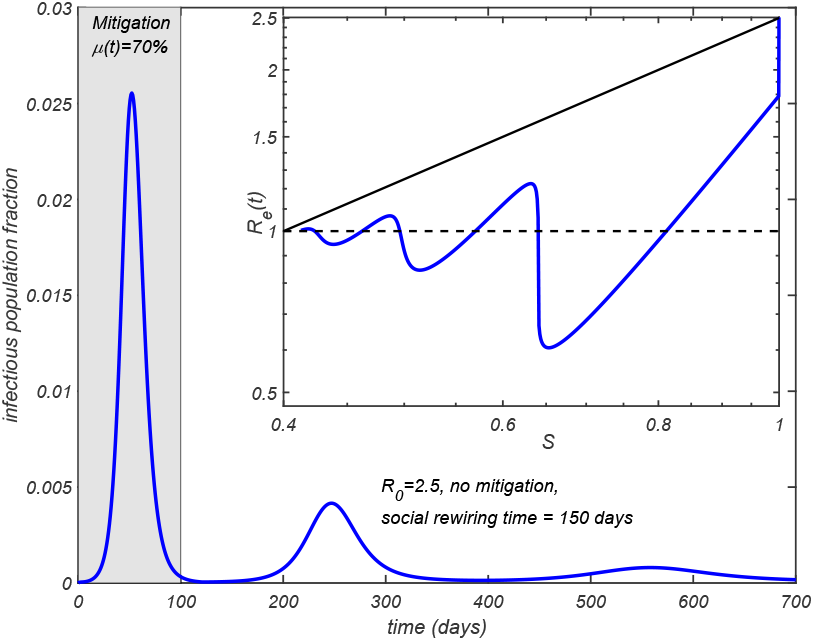
Effect of social rewiring on the epidemic dynamics. The time course of an epidemic in a heterogeneous SIR model with *R*_0_ = 2.5 and *λ* = 3. During the first 100 days a mitigation factor *µ* = 0.7 is applied. Social networks gradually rewire with a time constant *τ*_*s*_ = 150 days. The figure shows multiple waves. The inset shows *R*_*e*_(*t*) plotted as a function of *S*(*t*). Solid line shows the homogeneous limit reached after multiple waves.

Note that each of the waves follows the power law relationship between *R*_*e*_(*t*) ∼*S*(*t*)^*λ*^ predicted by Eq. 14. Since constant rewiring eliminates correlations in individual social activity on scales longer than *τ*_*s*_, the epidemic stops after multiple waves bring the total fraction of infected individuals close to the unmodified (homogeneous) herd immunity threshold 1*/R*_0_. Note, however that in this case there is almost no overshoot and thus the final size of the epidemic is reduced compared to the case of a purely homogeneous and unmitigated epidemic.

## Discussion

In this work, we have demonstrated how the interplay between short-term overdispersion and persistent heterogeneity in a population leads to dramatic changes in epidemic dynamics on multiple time scales, transient suppression of the epidemic during its early waves, all the way up to the state of long-term herd immunity. First, we developed a general approach that allows for the persistent heterogeneity to be easily integrated into a wide class of traditional epidemiological models in the form of two non-linear functions *R*_*e*_(*S*) and *S*_*e*_(*S*), both of which are fully determined by the statistics of individual susceptibilities and infectivities. Furthermore, *R*_*e*_(*S*) is largely defined by a single parameter, the immunity factor *λ*, introduced in our study. Like susceptibility itself, *λ* has two contributions: biological and social (see Eqs. (11-12)).

We then expanded our approach to include effects of time dependence of individual social activity, and in particular of likely correlations over the timescale of a single generation interval. While our results for purely persistent heterogeneity confirmed and corroborated that HIT would be suppressed compared to the homogeneous case, addition of temporal variations led to a dramatic revision of that simple narrative. Both persistent heterogeneity and short-term overdispersion contributions lead first to a slow down of a fast-paced epidemic, and to its medium-term stabilization. However, this state of Transient Collective Immunity (TCI) is fragile and does not constitute long-term herd immunity. HIT is indeed suppressed, but only due to the persistent heterogeneity. This suppression is significantly weaker than the initial stabilization responsible for the TCI state reached after the first wave of a fast-paced epidemic.

Among other implications of the TCI phenomenon is the suppression of the so-called overshoot. Namely, it is well known that most models predict that an epidemic will not stop once HIT is passed, ultimately reaching a significantly larger cumulative attack rate, FSE. Multiple prior studies (9, 10, 20, 21, 29, 33) have shown that FSE would be suppressed by persistent heterogeneity, similarly to HIT. In *SI Appendix*, we present a simple result that unifies several previously studied limiting cases, and gives an explicit equation for the FSE for the gamma-distributed susceptibility and variable level of its correlation with infectivity. However, because of the transient suppression of the early waves of the epidemic discussed in this work, the overshoot effect would be much weaker or essentially eliminated. For instance, our simple rewiring model demonstrates how the epidemic, after several waves, ultimately reaches HIT level, but does not progress much beyond it (see Fig. 5). The FSE result may still be used, but primarily as an estimate for the size of the first wave of an (unmitigated) epidemic. In that case, the transient value of immunity factor *λ*_eff_ should be assumed.

By applying our theory to the COVID-19 epidemic we found evidence that the hardest-hit areas such as New York City, have likely passed TCI threshold by the end of the first wave, but are less likely to have achieved a real long-term herd immunity. Other places that had intermediate exposure, such as e.g. Chicago, while still below the TCI threshold, have their effective reproduction number reduced by a significantly larger factor than predicted by traditional epidemiological models. This gives a better chance of suppressing the future waves of the epidemic in these locations by less disruptive measures than those used during the first wave, e.g. by using masks, social distancing, contact tracing, control of potential superspreading events, etc. However, similar to the case of NYC, transient stabilization of the COVID-19 epidemic in Chicago will eventually wane. As for the permanent HIT, although suppressed compared to classical value, it definitely has not yet been passed in those two locations.

In a recent study (34), the reduction of HIT due to heterogeneity has been illustrated using a toy model. In that model, 25% of the population was assumed to have their social activity reduced by 50% compared to a baseline, while another 25% had their social activity elevated twofold. The rest of the population was assigned the baseline level of activity. According to Eq. 12, the immunity factor in that model is *λ* = 1.54. For this immunity factor, Eq. (15) predicts HIT at *S*_0_ = 64%, 55% and 49%, for *R*_0_ = 2, 2.5, and 3, respectively. Despite the fact that the model distribution is not gamma-shaped, these values are in a very good agreement with the numerical results reported in Ref. (34): *S*_0_ = 62.5%, 53.5%, and 47.5%, respectively.

Thus there is a crucial distinction between the persistent heterogeneity, short-term variations correlated over the time scale of a single generation interval, and overdispersion in transmission statistics associated with short-term superspreading events (12, 13, 16, 26–28). In our theory, a personal decision to attend a large party or a meeting would only contribute to persistent heterogeneity if it represents a recurring behavioral pattern. On the other hand, superspreading events are shaped by short-time variations in individual infectivity (e.g. a person during highly infectious phase of the disease attending a large gathering). Hence, the level of heterogeneity inferred from the analysis of such events (12, 27) would be significantly exaggerated and should not be used to estimate the TCI threshold and HIT. Specifically, the statistics of superspreading events is commonly described by the negative binomial distribution with dispersion parameter *k* estimated to be about 0.1 for COVID-19 (28). According to Ref. (12), this is consistent with the expected value of the individual-level reproduction number *R*_*i*_ drawn from a gamma distribution with the shape parameter *k* ≃0.1. This distribution has a very high coefficient of variation, *CV* ^2^ = 1*/k*≃ 10. In the case of a perfect correlation between individual infectivity and susceptibility *α*, this would result in an unrealistically high estimate of the immunity factor: *λ* = 1+2*CV* ^2^ = 1+2*/k* ≃20. For this reason, according to our perspective and calculation, the final size of the COVID-19 epidemic may have been substantially underestimated in Ref. (30). Similarly, the degree of heterogeneity assumed in other recent studies (31, 48) is considerably larger than our estimates. Based on our analysis, the value of the immunity factor *λ* depends on the pace of the epidemic and on the timescale under consideration. We estimated its long-term value (responsible for the permanent HIT) as *λ*_∞_≈ 2. However, the transient values are expected to be higher, especially during the first several waves of COVID-19 in select locations, characterized by large growth rates. Our analysis of the empirical data in NYC and Chicago indicate that the slowdown of the epidemic dynamics in those locations was consistent with *λ*≈ 4. In Table 1 we present our estimates of the factor by which *R*_*e*_ is transiently suppressed as a result of depletion of susceptible population in selected locations in the world, as of early June 2020, as well as the predicted long-term suppression related to acquisition of a partial herd immunity.

**Table 1.** **Effects of heterogeneity on suppression of the effective reproduction number *R*_*e*_ in selected locations. The transient and long-term suppression coefficients *R*_*e*_ /*R*_*0*_ are calculated using *λ* = 4 and *λ* = 2, respectively. Fraction of susceptible population *S* as of early June 2020 is estimated from the cumulative reported death count per capita, assuming the infection fatality rate (IFR) of 0.7% (60).**

| Location | Deaths, per 1000 | Attack Rate (1-S) | $R_e$ suppression transient | $R_e$ suppression long-term |
| --- | --- | --- | --- | --- |
| New York City, USA (47) | 2.1 | 30% | 0.24 | 0.50 |
| Lombardy, Italy (61) | 1.7 | 24% | 0.33 | 0.58 |
| London, UK (62) | 0.9 | 13% | 0.58 | 0.75 |
| Chicago, USA (54) | 0.9 | 13% | 0.58 | 0.75 |
| Stockholm, Sweden (63) | 0.9 | 13% | 0.58 | 0.75 |

Population heterogeneity manifests itself at multiple scales. At the most coarse-grained level, individual cities or even countries can be assigned some level of susceptibility and infectivity, which inevitably vary from one location to another reflecting differences in population density and its connectivity to other regions. Such spatial heterogeneity will result in self-limiting epidemic dynamics at the global scale. For instance, hard-hit hubs of the global transportation network such as New York City during the COVID-19 epidemic would gain full or partial TCI thereby limiting the spread of infection to other regions during future waves of the epidemic. This might be a general mechanism that ultimately limits the scale of many pandemics, from the Black Death to the 1918 influenza.

## Data Availability

The manuscript uses data provided by the Illinois Department of Public Health through a Data Use Agreement with Civis Analytics. The source code for the model is freely available online at https://github.com/uiuc-covid19-modeling/pydemic
https://github.com/uiuc-covid19-modeling/pydemic

## ACKNOWLEDGMENTS

We gratefully acknowledge discussions with Mark Johnson at Carle Hospital. The calculations we have performed would have been impossible without the data kindly provided by the Illinois Department of Public Health through a Data Use Agreement with Civis Analytics. This work was supported by the University of Illinois System Office, the Office of the Vice-Chancellor for Research and Innovation, the Grainger College of Engineering, and the Department of Physics at the University of Illinois at Urbana-Champaign. Z.J.W. is supported in part by the United States Department of Energy Computational Science Graduate Fellowship, provided under Grant No. DE-FG02-97ER25308. A.E. acknowledges partial support by NSF CAREER Award No. 1753249. This work made use of the Illinois Campus Cluster, a computing resource that is operated by the Illinois Campus Cluster Program (ICCP) in conjunction with the National Center for Supercomputing Applications (NCSA) and which is supported by funds from the University of Illinois at Urbana-Champaign. This research was partially done at, and used resources of the Center for Functional Nanomaterials, which is a U.S. DOE Office of Science Facility, at Brookhaven National Laboratory under Contract No. DE-SC0012704.

## Supporting Information Appendix

### Derivation of quasi-homogeneous model

#### Age-of-infection model

We start we the same age-of-infection model as described in the main text, but include additional time-dependent modulation of the force of infection :

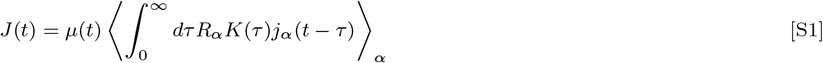

Here, the modulation factor *µ*(*t*) can be due (e.g.) to mitigation measures or seasonal forcing. Due to this modification, Eq. (5) should be rewritten as follows:

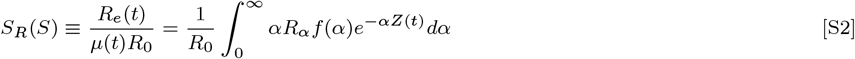

Here 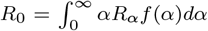is the basic reproduction number. Now one can write an integral equation for force of infection which is formally identical to the one for a homogeneous case:

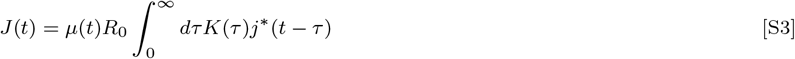

Here introducing infectivity-weighted incidence rate, *j*^*∗*^ = *S*_*R*_*J*. Eq. (7) completes the set of our quasi-homogeneous equations:

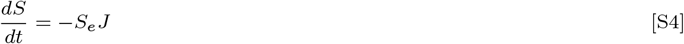

As discussed in the main text, the inhomogeneity is fully accounted for by non-liner function *S*_*R*_(*S*), and variable effective susceptibility *α*_*e*_(*S*).

#### Compartmentalized SIR/SEIR models

The basic SIR and SIER models can be viewed as particular cases of the age-of infection model discussed above. However, because of their great importance and wide use, we present our construction for a specific case of SEIR:

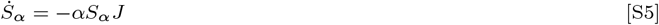

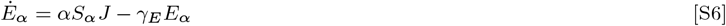

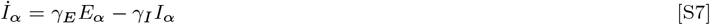

Here, the force of infection is 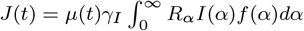. We define infectivity-weighted “Exposed” and “Infectious” fractions as

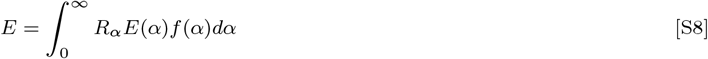

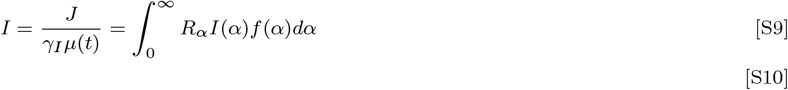

This leads to a complete description of epidemic dynamics with three ordinary differential equations, formally equivalent to those for the homogeneous case. The difference are, once again, functions *R*_*e*_ = *µ*(*t*)*S*_*R*_(*S*)*R*_0_ and *S*_*e*_(*S*):

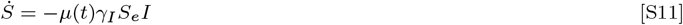

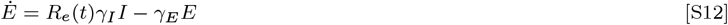

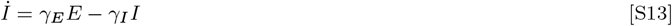

#### Correlation parameter and scaling relationship between infectivity and susceptibility

Below we consider a model in which biological susceptibility *α*_*b*_ is correlated neither with infectivity nor with social strength *α*_*s*_ of an individual. On the other hand, both the overall susceptibility and infectivity are proportional to *α*_*s*_. Let *f*_*x*_ and *f*_*y*_ be probability density functions (pdfs) of variables *x*≡ ln *α*_*s*_ and *y* ≡ ln *α*_b_. It is reasonable to assume a log-normal distribution for *α*_b_, since biological susceptibility can be modeled as a product of several random factors (due to age, gender, genetics, pre-existent conditions, etc). This corresponds to a Gaussian form for *f*_*y*_ with variance *σ*^2^ and mean −*σ*^2^*/*2 (assuming normalization ⟨*α*_*b*_⟩ = 1). For a given value of *α*, this translates into Gaussian distribution of variable *x* with the same variance, and mean ln *α* + *σ*^2^*/*2. This allows us to calculate the average *α*_*s*_ which is proportional to *R*_*α*_:

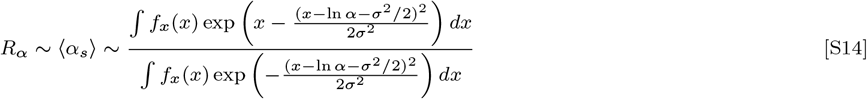

This integral can be evaluated by the method of steepest descents: for most pdfs *f*_*x*_ and *f*_*y*_, will be dominated by the vicinity of point *x*_0_ defined by the condition *f*′(*x*_0_)*/f* (*x*_0_) = (*x*_0_*/σ*^2^− 1*/*2). By expanding ln *f* (*x*) in *x*′ = *x− x*_0_, we obtain *f*_*x*_ (*x*′)≈ *f*(*x*_*σ*_) exp(*rx′ − κx*′^2^*/*2), where *r* = *f*′(*x*_0_)*/f* (*x*_0_) = *x*_0_*/σ*^2^−1*/*2 and *κ* = −*f*″ (*x*_0_)*/f* (*x*_0_) + *r*^2^. After substituting this Gaussian approximation for *f*_*x*_ back into the above equation, we obtain the scaling relationship between *α* and *R*_*α*_

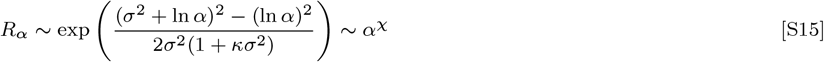

Here *χ* = 1*/*(1 + *κσ*^2^).

**Functions** *S*_*R*_(*S*) **and** *S*_*e*_(*S*). According to Eq.(4), function *S*(*Z*) is directly related to the moment generating function *M*_*α*_ for pdf *f* (*α*)

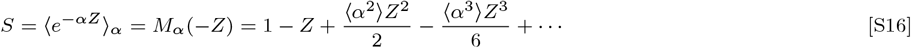

This function also determines the effective fraction of susceptible population *S*_*e*_:

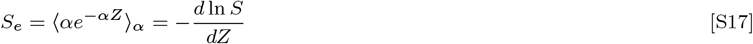

Remarkably, once the function *S*_*e*_(*S*) is found, it completely determines how *S*_*R*_, and hence *R*_*e*_, behaves in the limiting cases of both the strong and weak correlations:

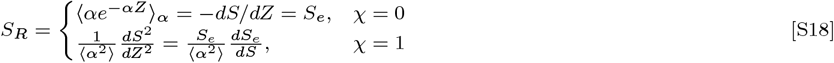

### Application to specific distributions of susceptibility

#### Gamma distribution

Consider the gamma distribution with ⟨*α*⟩ = 1 and 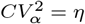:

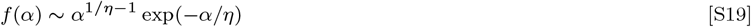

By using Eqs. (4)-(5), we obtain:

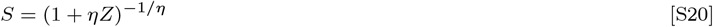

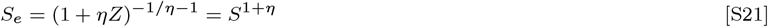

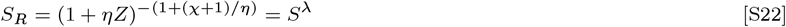

This leads to the scaling relationship *R*_*e*_ = *R*_0_*S*^*λ*^, Eq. (14).

#### Truncated power law distribution

We now consider power law distributed *α, f* (*α*) ∼1*/α*^1+*s*^ (*s >* 0), with upper and lower cut-offs, *Eα*_+_ and *α*_+_, respectively. If the upper cut-off is implemented as an exponential factor exp(− *α/α*_+_), we recover the functional form identical to the gamma distribution, Eq. (S19) discussed above, but with negative values of the shape factor:

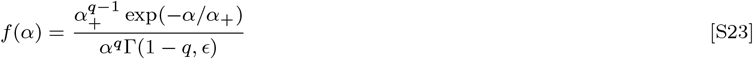

Due to the normalization ⟨*α*⟩ = 1,

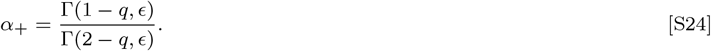

In the case of gamma distribution, the coefficient of variation *CV*_*α*_ would completely determine the overall shape of pdf. For power law with exponent 1 ≤ *q* ≤ 3, the value of *η* = *CV* ^2^ sets the dynamic range the between upper and lower cut-offs, i.e. the parameter *ϵ*:

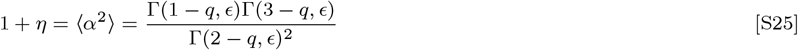

By using Eq. (4)-(5), we can obtain exact results for *S S*_*R*_ in terms of *Z*:

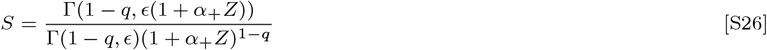

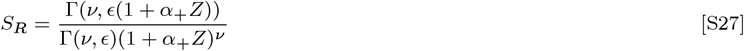

Here *ν* = 2 + *χ* − *q*. The resulting function *R*_*e*_*/R*_0_ = *S*_*R*_(*S*) is shown in Fig. S1 for several values of the exponent *q*.

For *χ* = 0, the overall function *S*_*R*_(*S*) = *S*_*e*_(*S*) can be very well fitted by an empirical analytic formula that depends only on 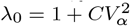 and an additional shape parameter *Δ*_*λ*_ = *CV*_*α*_(*γ*_*α*_ − 2*CV*_*α*_):

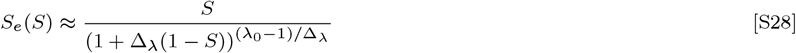

According to Eq. (S18), this function completely defines behavior of *S*_*R*_ in both limits of the weak and strong correlation regimes :

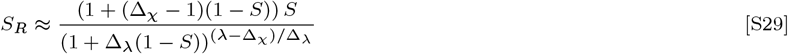

Here *Δ*_*χ*_ = (*Δ*_*λ*_ + 1)*/λ*_0_, and *λ* = *λ*_1_ for *χ* = 1. For *χ* = 0, *δ*_*χ*_ has to be set to 1.

#### Log-normal distribution

The log-normal distribution is a very natural candidate to describe statistics of *α*. It universally emerges for multiplicative random processes. Transmission of an infection involves a complex chain of random events, both social and biological, which can be conceptualized as such multiplicative process. For instance, it may depend on how likely a given person would be involved in a potential superspreading event, how likely that person would have a close contact with a potential infector, what would be the duration of their contact, how effective the individual immune system is in preventing and suppressing the infection.

For the log-normal distribution, the initial drop in *R*_*e*_ according to Eq. (10), is noticeably faster than for a gamma distribution: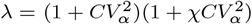.However, the initial linear regime is also much narrower. Figure S1 shows the dependence *R*_*e*_(*S*) for the log-normal distribution alongside with the above results for gamma and power law distributions computed for the same values of CV (specifically,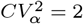. As one can see from these plots, despite a stronger effect of heterogeneity at the early stage, the curves generated by log-normal distribution approach *R*_*e*_ = 0 significantly slower than those corresponding to the gamma distribution. Note that the overall behavior of *R*_*e*_(*S*) generated by the log-normal distribution closely matches the one obtained for the power law distribution with a certain scaling exponent *q*. This exponent would depend on *CV* and should approach 1 in the limit of sufficiently wide distribution when the log-normal pdf asymptotically approaches a power law 1*/α* with upper and lower cut-offs.

### Final Size of Epidemic

Here we derive a simple result for FSE in a population with a persistent heterogeneity. To do this, we integrate Eq. (6) over time *t*. This yields a relation 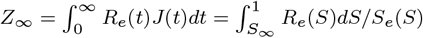 for the final value of *Z* when the epidemic has run its course, and this in turn can conveniently be expressed in terms of the fraction of the susceptible population, *S*_*∞*_:

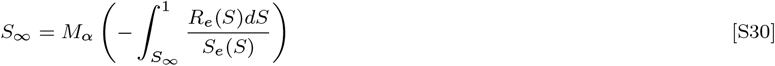

This equation is valid for an arbitrary distribution of *α*, arbitrary correlation between susceptibility and infectivity, and for any statistics of the generation interval. This result can be also obtained as a solution to a general integral equation derived in Ref. (21) for the well-mixed case. Eq. S30 combines and generalizes several well-known results: (i) in the weak correlation limit (*R*_*α*_ = *R*_0_), when the integral in the r.h.s. is equal to *R*_0_(1− *S*_∞_), Eq.(S30) reproduces results of Refs. (20, 21, 29, 33), (ii) in the opposite limit of a strong correlation (*R*_*α*_∼*α*), the integration gives *R*_0_(1 −*S*_*e*_(*S* _∞_))*/ ⟨α*^2^⟩, and one recovers the result for the FSE on a network (9, 10, 21).

For the case of gamma-distributed persistent susceptibility Eq. (S30), gives:

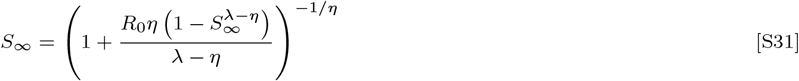

It should be emphasized however that this result is of limited relevance to more realistic situations. Even if one assumes no government-imposed mitigation or societal response to the epidemic, the case of fully persistent heterogeneity is just an approximation. As we demonstrate in our paper, short-term correlations of time-dependent individual susceptibilities and infectivities lead to transient stabilization of a fast-pacing epidemic. Because of this effect, Eq.(S30)-Eq.(S31) should be interpreted as an estimate of the size of the first wave rather than the actual FSE.

### Path-integral theory of epidemic with time-dependent heterogeneity

Here we present a generalization of the theory developed in the previous section that incorporates the effects of time variations of individual susceptibilities and infectivities, as well as temporal correlations between them. Since these fast variations are primarily caused by bursty dynamics of social interactions, and since heterogeneous biological susceptibility appears subdominant in the context of COVID-19, we set *α*_*b*_ = 1 for all individuals, so that *α* has purely social origin. Let *a*_*i*_(*t*) = *α*_*i*_ + *δa*_*i*_(*t*) be the time-dependent susceptibility of a person. Because of the social nature of *a*(*t*), one’s individual infectivity is also proportional to it at any given time: *β*_*i*_(*t*) = *R · K*(*τ*)*a*_*i*_(*t*). As before, *τ* is time from infection, *K*(*τ*) is the pdf of generation intervals. Accordingly *R* is individual reproductive number of an “average” person with social activity *a*_*i*_(*t*) = 1, in the fully susceptible population. The state of an individual is described by a step function *s*_*i*_(*t*) which is 1 as long as the person is susceptible, and turns to 0 at the moment of infection. The time evolution of the epidemic follows a stochastic generalization of Eqs. (1)-(2):

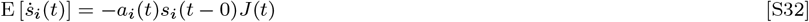

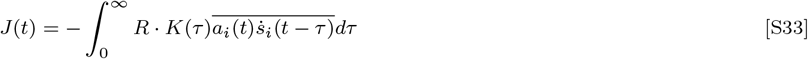

Here bar 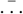 represents averaging over individual members of population (indexed by *i*), in contrast with ⟨…⟩, averaging over all subgroups with various values of persistent heterogeneity *α*. E[…] stands for expected value.

The overall quasi-homogeneous description given by Eqs.(7)-(6), remains valid. It is obtained by averaging Eqs. (S32)-(S33) over the entire population. However, in contrast to the case of persistent heterogeneity, variables *S*(*t*), *S*_*e*_(*t*) and *R*_*e*_ are no longer connected to each other by a simple functional relationship. To relate them we first note that the average probability that an individual is still susceptible at time *t* is given by 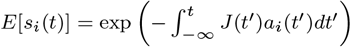 Therefore,

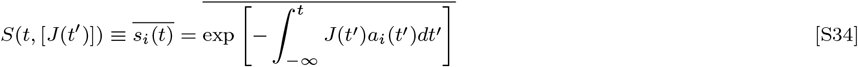

In other words, *S* becomes a functional over the set of all possible epidemic trajectories *J* (*t*). It still has the structure of a moment generating function for the field *a*_*i*_(*t*), and thus is a direct analogue of the partition function broadly used in statistical physics, stochastic calculus, and field theory. The specific form of this functional depends on probabilities assigned to different individual trajectories *a*_*i*_(*t*). As a natural generalization of the case of persistent heterogeneity, *S*_*e*_(*t*) and *R*_*e*_(*t*) can be obtained as, respectively, the first and the second variations of the functional *S* over *J* (*t*):

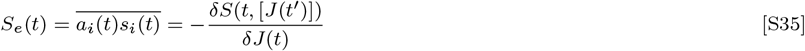

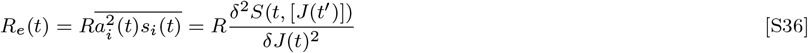

For the sake of simplicity, in deriving Eq.(S36) we assumed *α*_*i*_(*t*) to be smoothed over the timescale of a single generation interval. As a result,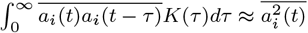. By applying Eq.(S36) to the initial state of fully susceptible population we obtain the result for *R*_0,_Eq.(16)

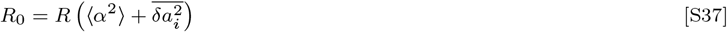

At the early stages of epidemic 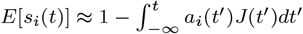 for the entire population. After substituting this expression for *s*_*i*_ (*t*) to Eq.(S36) one obtains a generalization of our previous result for the initial suppression of *R*_*e*_, Eqs.(9)-(10):

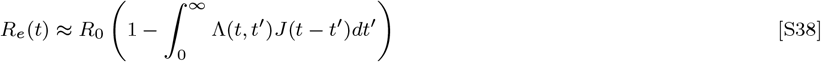

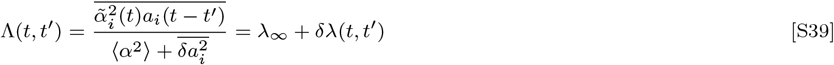

Here *λ* _∞_= Λ(∞) and *δλ*(*t, t*^*‘*^) are the constant and time-dependent contributions to “immunity kernel” Λ(*t, t*^*‘*^) which are discussed in the main text.

To obtain a corrected result for HIT, we assume a very slow progression of the epidemic (e.g. due to a gradual relaxation of the level of mitigation). In this case, any intermediate-term correlations between time dependent variations *δα*_*i*_(*t*) become negligible, and we largely recover the formalism developed for pure persistent heterogeneity. Under the same assumption that was used for the estimate of *λ*_*∞*_ in the main text (i.e.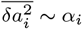), the only modification that needs to be done is to replace the term *αR*_*α*_ in Eq. (5) with (*χ*^*∗*^*α*^2^ + (1 − *χ*^*∗*^)*α*)*R*_0_(where 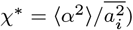. This gives an expression for *R*_*e*_(*S*) in terms of two earlier results, 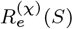 (for *χ* = 0 and 1, respectively):

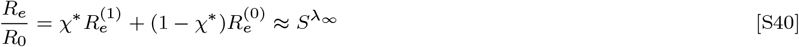

Here *λ*_∞_ = 1 + (1 + *χ*^*∗*^)*η*. To obtain this result, we substituted the scaling functions for gamma-distributed persistent heterogeneity, *R*_*e*_(*S*) = *R*_0_*S*^*λ*^, with *λ* = 1 + (1 + *χ*)*η*, as given by Eq.(14).

**Fig. S1.**
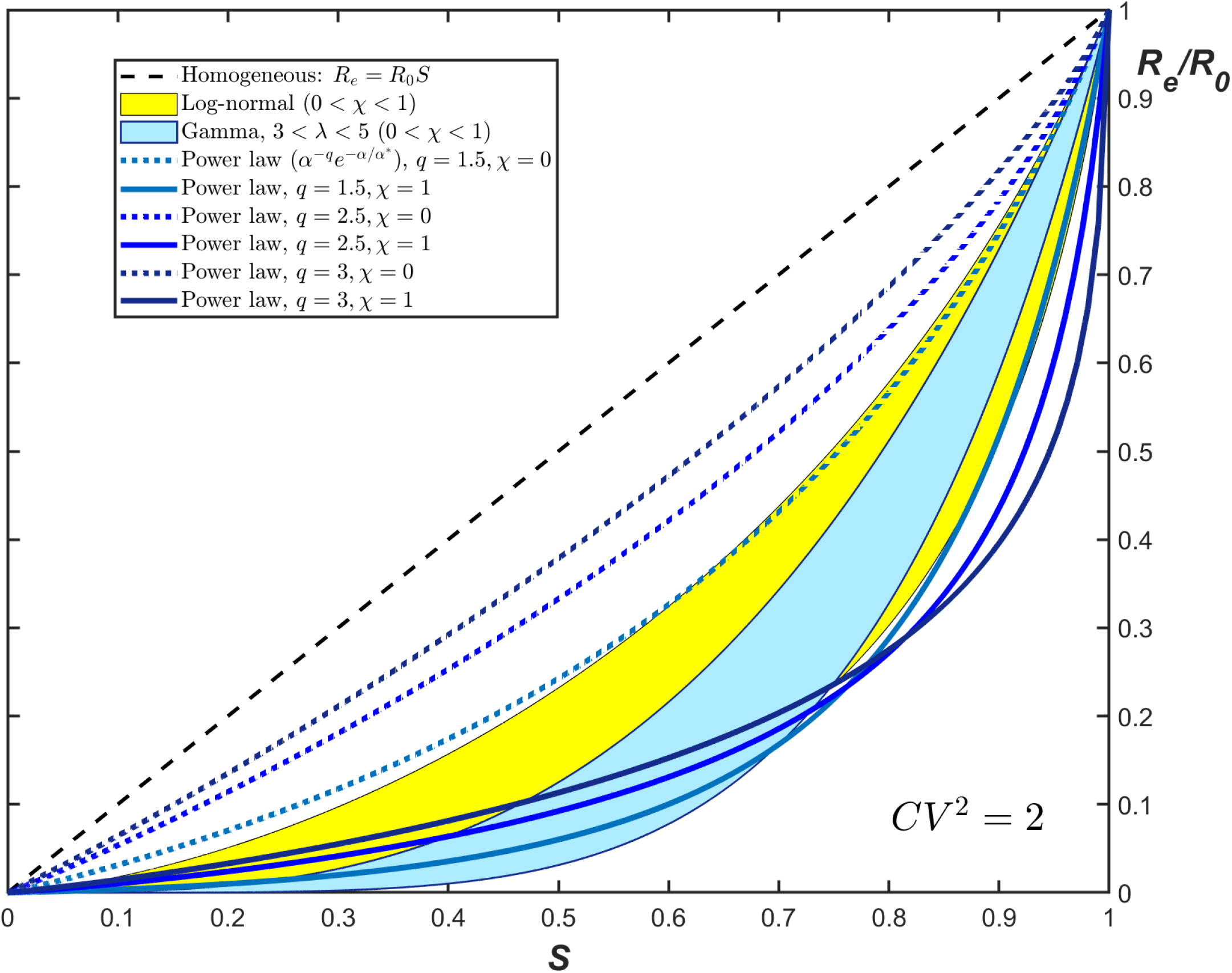
*R*_*e*_*/R*_0_ vs *S* dependence for three different families of probability distribution *f* (*α*): Gamma (light blue), truncated power law (dashed lines), and log-normal (yellow). Different curves correspond to the same value of the coefficient of variation 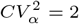, and two limiting values (0 and 1) of the correlation parameter *χ*.

**Fig. S2.**
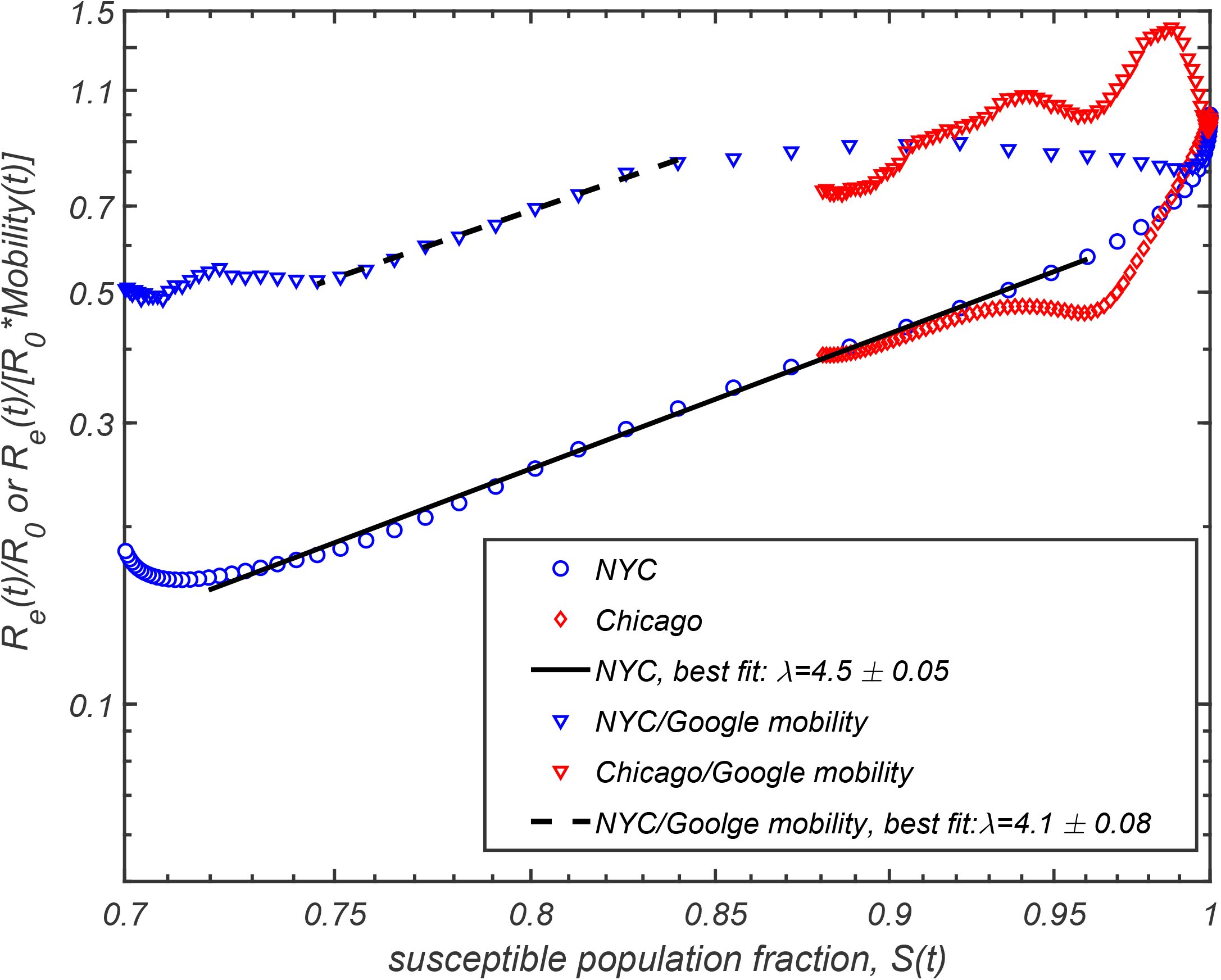
Exploration of effect of mobility on data presented in Figure 3(A). Triangles represent data points for NYC and Chicago with *R*_*e*_(*t*)*/R*_0_ corrected by a mobility factor calculated from Google community mobility report, Ref. (57). We compute the mobility for NYC using average mobility of its five counties: New York county, Bronx county, Kings county, Richmond county, and Queens county, weighted by their population fraction.

**Fig. S3.**
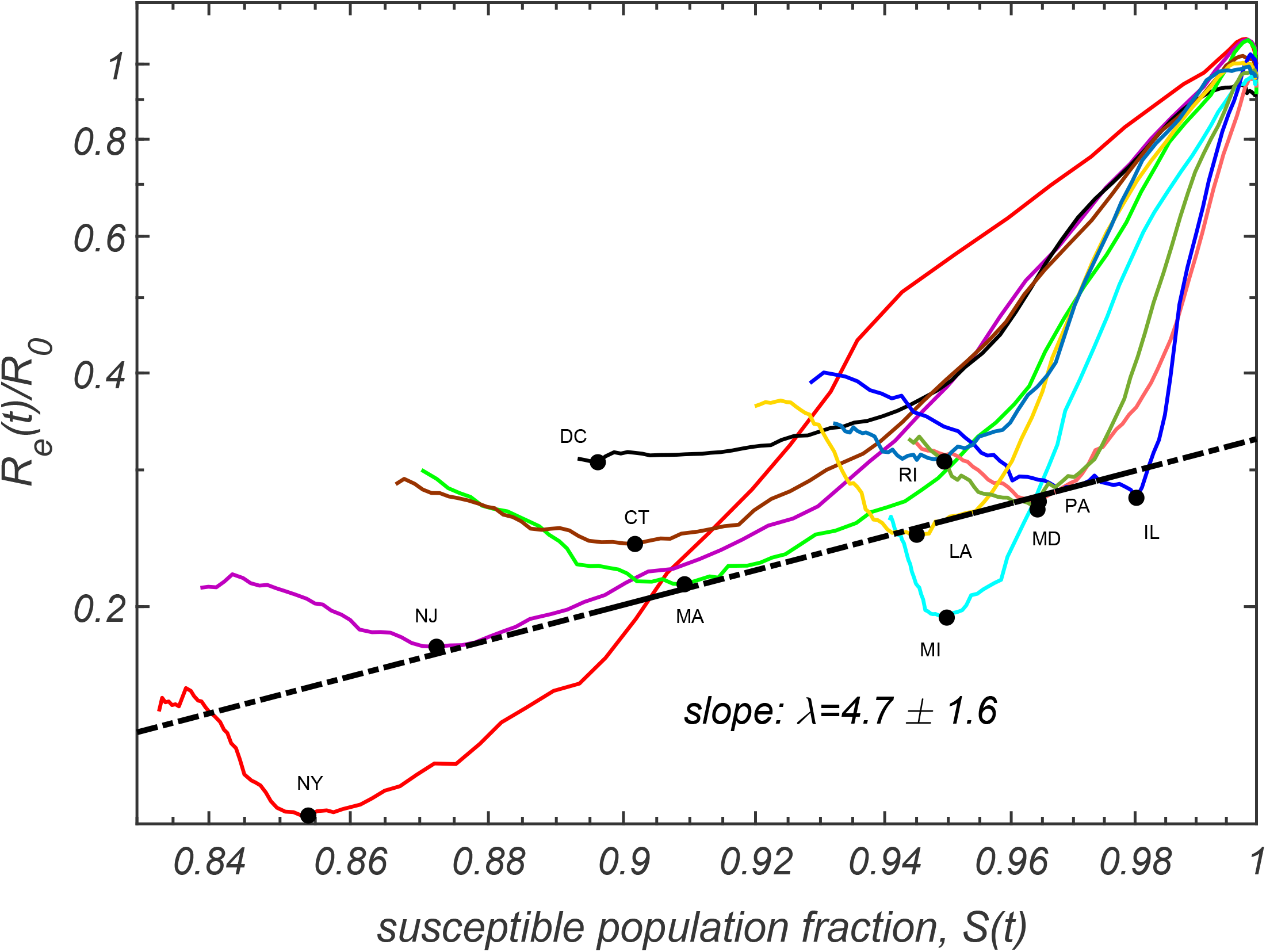
Time progressions of *Re*(*t*)*/R*_0_ and *S*(*t*) for the hardest-hit US states and DC, as reported in Ref. (46). Black dots correspond to absolute minima of transmission and population susceptible fractions. The dashed line with slope *λ* = 4.7 *±* 1.6 is the best power law fit through these black dots.

**Fig. S4.**
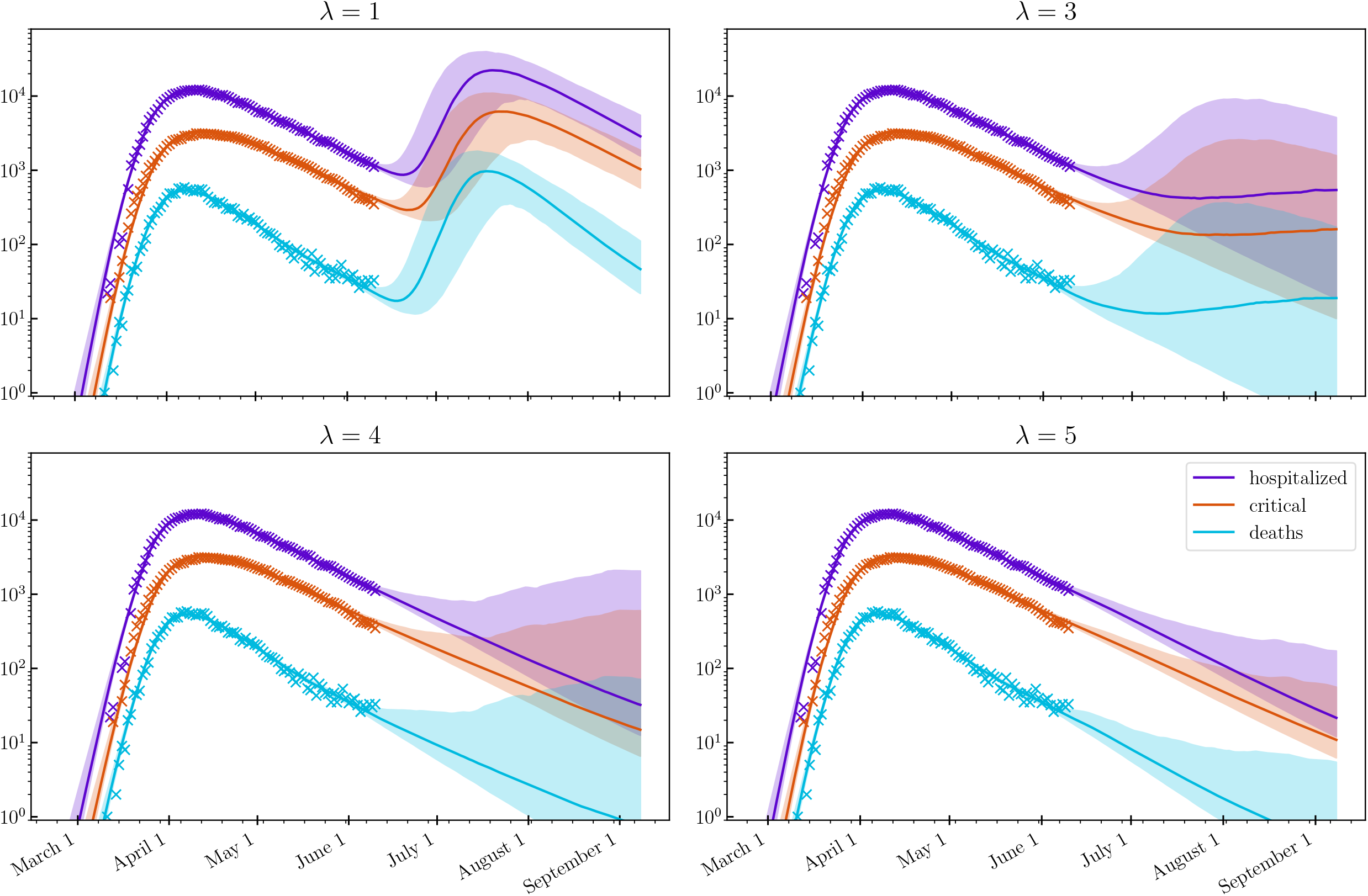
Hospitalization, ICU occupancy and daily deaths in NYC modeled under hypothetical scenario when any mitigation is completely eliminated as of Jun 15 2020, for various values of *λ*. Model described in Ref. (40) is calibrated on data from Ref.(54), up to June 10, 2020 (shown as crosses). 95% confidence intervals are indicated.

**Fig. S5.**
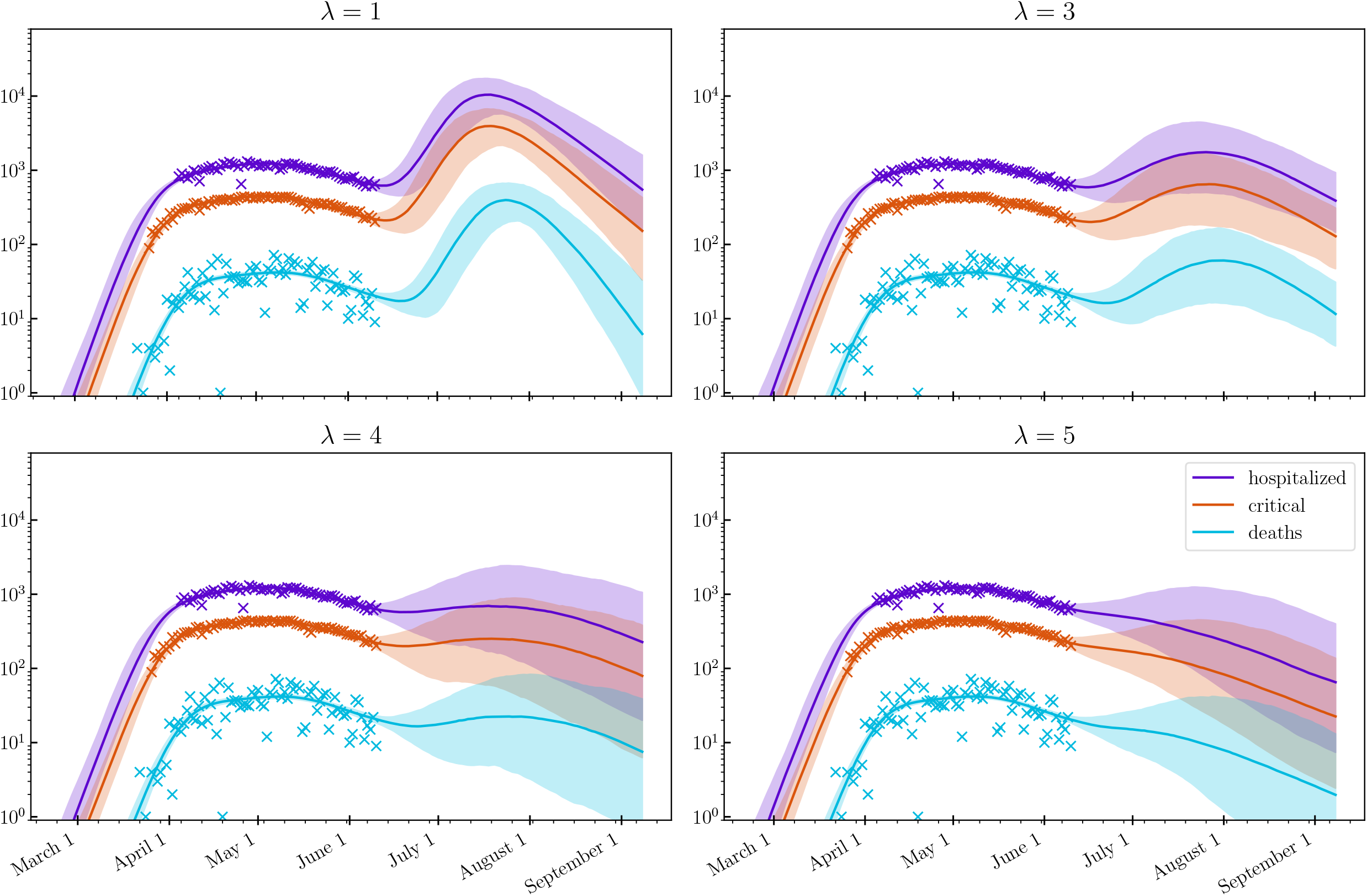
Hospitalization, ICU occupancy and daily deaths in Chicago modeled under hypothetical scenario when any mitigation is completely eliminated as of Jun 15 2020, for various values of *λ*. Model described in Ref. (40) is calibrated on data from Ref.(54), up to June 10, 2020 (shown as crosses). 95% confidence intervals are indicated.

## Notes

### Competing Interest Statement

The authors have declared no competing interest.

### Funding Statement

Our calculations would have been impossible without the data kindly provided by the Illinois Department of Public Health through a Data Use Agreement with Civis Analytics.  This work was supported by the University of Illinois System Office, the Office of the Vice-Chancellor for Research and Innovation, the Grainger College of Engineering, and the Department of Physics at the University of Illinois at Urbana-Champaign. Z.J.W. is supported in part by the United States Department of Energy Computational Science Graduate Fellowship, provided under Award No. DE-FG02-97ER25308. This work made use of the Illinois Campus Cluster, a computing resource that is operated by the Illinois Campus Cluster Program (ICCP) in conjunction with the National Center for Supercomputing Applications (NCSA) and which is supported by funds from the University of Illinois at Urbana-Champaign. This research was partially done at, and used resources of the Center for Functional Nanomaterials, which is a U.S. DOE Office of Science Facility, at Brookhaven National Laboratory under Contract No.~DE-SC0012704.

### Author Declarations

This manuscript does not involve research on human subjects. The public data used in this study contains no identifiable private information.

### Summary of Updates

Minor revisions

